# Enhancing Pandemic Prediction: A Deep Learning Approach Using Transformer Neural Networks and Multi-Source Data Fusion for Infectious Disease Forecasting

**DOI:** 10.1101/2025.06.24.25330211

**Authors:** Jiande Wu, Shakhawat Tanim, MinJae Woo, Tanvir Ahammed, Lior Rennert

## Abstract

The Covid-19 pandemic has highlighted the urgent need for accurate prediction of pandemic trends. We propose a deep learning model for predicting Covid-19 cases and deaths at the county level through transformer neural networks with multi-source data fusion, incorporating historical case data, death data, and social media sentiment analysis to capture both temporal (historical trends) and spatial (geographical relationships) dynamics within time series data. Additionally, we develop multi-level and multi-scale attention mechanisms for adaptive time-frequency analysis. Across three Omicron variant waves (December 2021 through February 2023), the model demonstrated strong performance in predicting county-level Covid-19 cases and deaths, with median county agreement accuracy ranging from 74.0% to 82.6% for one-week case forecasts and 68.7% to 79.6% for 5-week case forecasts. Median county agreement accuracy for deaths ranged from 83.2% to 86.3% for one-week forecasts and 84.3% to 87.2% for five-week forecasts. Incorporating social media data yielded mild to moderate improvement in forecasting accuracy. Overall, the proposed model yielded substantial improvements compared to a baseline persistence model utilizing the last observation carried forward. By integrating real-time data and capturing complex pandemic dynamics, this approach surpasses traditional methods. Its high accuracy and generalizability make it a valuable tool for enhancing public health preparedness and response strategies in future outbreaks.

## 1 Introduction

The coronavirus disease 2019 (Covid-19) pandemic has posed unprecedented challenges to public health systems around the world. The rapid spread of the virus has put pressure on medical facilities and tested the resilience of medical professionals and institutions [1]. Coordinating response efforts, implementing preventive measures, and ensuring medical resource availability are critical tasks for governments and medical institutions [2]. The complex and ever-evolving challenges posed by the pandemic, demand creative solutions and flexible response strategies [3]. The pandemic exposed healthcare system vulnerabilities and the urgency for advance outbreak prediction. Predictive modeling is key to informing preparedness and response [4], [5]. This underscores the significance of ongoing research and collaborative efforts to enhance our understanding of infectious diseases and improve our ability to forecast and mitigate their impact.

Deep learning methods have revolutionized predictive modeling, achieving superior performance across various forecasting tasks [6]. Their ability to identify complex patterns in vast datasets makes them well-suited for fields like natural language processing, image recognition, and financial forecasting [7]. This capability extends to pandemic forecasting [8]. As pandemic data, which encompasses factors like human behavior, healthcare capacity, and viral mutations, is inherently complex and dynamic, it aligns perfectly with the strengths of deep learning methods. Deep learning models, including recurrent neural networks (RNNs), long short-term memory networks (LSTMs), and attention-based transformers, can capture these complicated patterns, leading to more accurate predictions and better decision-making [9]. As deep learning advances, its integration into pandemic forecasting holds immense promise for improving our ability to anticipate and effectively respond to future public health threats.

Social media platforms like X (formerly Twitter) are valuable data sources, offering insights into public sentiment and behavior during pandemics [10]. Analyzing this data can significantly improve forecasting accuracy and timeliness [11]. Social media reveals early signs of emerging trends, concerns, and sentiment around the crisis [12]. Importantly, social media data can overcome limitations of hospital encounter data, which is often used to forecast disease trends but is not representative of all populations due to disparities in health care access [13]. By tracking user-generated content, researchers can identify potential outbreaks and understand public response to the pandemic [14]. This data, especially when integrated with deep learning models, allows us to capture subtle patterns that might influence the course of the pandemic [15]. As social media continues to evolve, leveraging its real-time insights is crucial for refining pandemic predictions and informing public health interventions.

Current epidemic forecasting methods often only depend on delayed indicators like confirmed cases [16]. This research proposes a new approach to overcome this limitation by incorporating diverse data sources. Social media data can provide immediate insight on disease dynamics, including infection rates, geographical distribution, and severity of illness. [17]. Researchers have found that social media can reveal early signs of emerging trends before local outbreaks [18]. By leveraging internet-based digital traces like Google searches and Twitter data, these researchers developed methods to anticipate sharp increases in Covid-19 activity, effectively detecting potential outbreaks earlier than traditional surveillance systems. This approach focused on early detection rather than precise case forecasting. While research has shown promise in using social media to detect early signs of emerging trends before local outbreaks, limitations exist. For instance, previous studies often focused on county-level analyses in a subset of highly populated counties (over 1 million inhabitants) [18]. This approach might not capture the nuances of outbreaks in less populous areas. Additionally, important community-level information is excluded from these analyses.

Our model advances county-level COVID-19 prediction beyond prior work like COURAGE [19] and the County Augmented Transformer [20] through key innovations. While sharing a focus on county-level forecasting, attention mechanisms, and external data with COURAGE, our model incorporates more granular features, a hybrid architecture blending attention with a state-specific training module. Compared to the County Augmented Transformer, which also utilizes attention and external data for spatial-temporal hospitalization modeling, our approach emphasizes state-specific adaptation for enhanced accuracy in South Carolina alongside generalizability, further integrating a wider array of data sources, including social media, to capture nuanced COVID-19 trends.

This study aims to develop and implement a deep learning model based on social media data, historical Covid-19 cases and deaths, and community-level contextual factors to forecast Covid-19 trends in South Carolina (SC) counties. This approach aims to provide a more comprehensive understanding of how social media can inform infectious disease forecasts across different population patterns. We leverage data from all counties within the state and incorporate demographic data to achieve a more comprehensive understanding of how social media can inform infectious disease forecasting across varying population landscapes. We develop a deep learning model integrating these diverse sources to capture the complex factors influencing Covid-19 outbreaks. This has the potential to predict outbreaks earlier, pinpoint high-risk areas, and inform targeted interventions [21], ultimately improving public health preparedness and response.

## 2 Materials and Methods

In our study, we collected multiple data sources for 46 South Carolina counties between January 2020 and March 2023.

### 2.1 Data Sources

In this section, we present and elucidate the epidemiological reporting of Covid-19, along with health-related searches and discussions pertaining to Covid-19 on Twitter.

To gain a comprehensive understanding of the Covid-19 landscape, our analysis relies on data from two trusted sources. These include The New York Times [22]. Their data is aggregated from state and local health agencies across the US, providing valuable insights at the county level. Specifically, we utilize data on confirmed and probable Covid-19 cases (cumulative and daily) and Covid-19 deaths (cumulative and daily) for South Carolina counties. We also utilize data from the Centers for Disease Control and Prevention (CDC) website [23]. The data points from the CDC include positive cases, hospitalization rates, and testing data. We conducted a comprehensive analysis of Covid-19 activity across 46 counties in the U.S. state of South Carolina spanning from January 1, 2020, to March 1, 2023. The data was collected daily and has 10 columns. The ‘cases 7-day average’ and ‘deaths 7-day average’ are the target variable. All other variables (along with time) were used as input features. The column names and description are specified in Table 1. This combined dataset offers a rich variety of metrics, including:

- Cases per 100,000 population (7-day change)
- Percent positive test results (last 7 days)
- Covid-19 deaths
- Total deaths
- Population by health service area
- Average cases per 100,000 population
- Average deaths per 100,000 population

**Table 1.** Description of variables in the dataset.

| Variable | Description | Comments |
| --- | --- | --- |
| date | Date of data release |  |
| county | County name |  |
| percent_test_results_reported_positive_last_7_days | Percentage of positive diagnostic and screening nucleic acid amplification tests (NAAT) during the last 7 days |  |
| covid_hospital_admissions_per_100k 7-day total | New Covid-19 admissions per 100,000 population (7-day total) |  |
| percent covid_inpatient_bed_utilization 7-day average | Percent of staffed inpatient beds occupied by Covid-19 patients (7-day average) |  |
| cases 7-day average | New Covid-19 (7-day average) | Target |
| deaths 7-day average | Deaths involving Covid-19, coded to ICD-code U07.1 |  |
| Total Deaths 7-day average | Deaths from all causes of death | Additional input features |
| county_population | County population (2019 Census estimate) |  |
| health_service_area_population | Health Service Area population (2019 Census estimate) * |  |

\* Health Service Areas (HSAs) are geographic regions defined by the National Center for Health Statistics (NCHS) to group counties with a shared focus on hospital care.

By incorporating these diverse metrics, our analysis can capture the nuances of the Covid-19 situation.

To strengthen the context of our analysis, we incorporate county-level census data from the U.S. Census Bureau [24]. This data variables include the overall county population and the population of each health service area. This enrichment layer provides demographic details and population statistics, offering a more comprehensive understanding of the factors at play.

For Twitter data related to Covid-19, our collection process is facilitated through the Sprinklr website [25]. Employing geolocation data, sentiments and targeted keywords, we filter relevant information within a specific timeframe. Sentiment labels are sourced from Sprinklr’s pre-processed outputs, with no additional text encoding. This approach allows us to extract real-time insights from social media, capturing public sentiments, emerging trends, and discussions surrounding the ongoing pandemic. All twitter used in this study were in SC. To filter the tweet data related to Covid-19, we used a simple keyword list, as shown in Table *2*. The list includes General Covid-19 terms, such as “covid,” “covid19,” “covid-19,” and “coronavirus.” Keywords related to vaccines, including “vaccine,” “effects AND vaccine,” and “side effects AND vaccine.” Terms related to Covid-19 symptoms, like “symptoms,” “Abdominal AND pain,” “Acute AND bronchitis,” “Anosmia,” and “Anxiety.” We applied sentiment filtering and temporal alignment to enhance the quality of the Twitter data and reduce the influence of unrelated or politicized content. Sentiment filtering was conducted using the keyword to isolate tweets related to health behaviors and Covid–19 relevant discussions, excluding political discourse. Temporal alignment was performed by synchronizing Twitter-derived sentiment trends with confirmed case and death data at the county level.

**Table 2.**
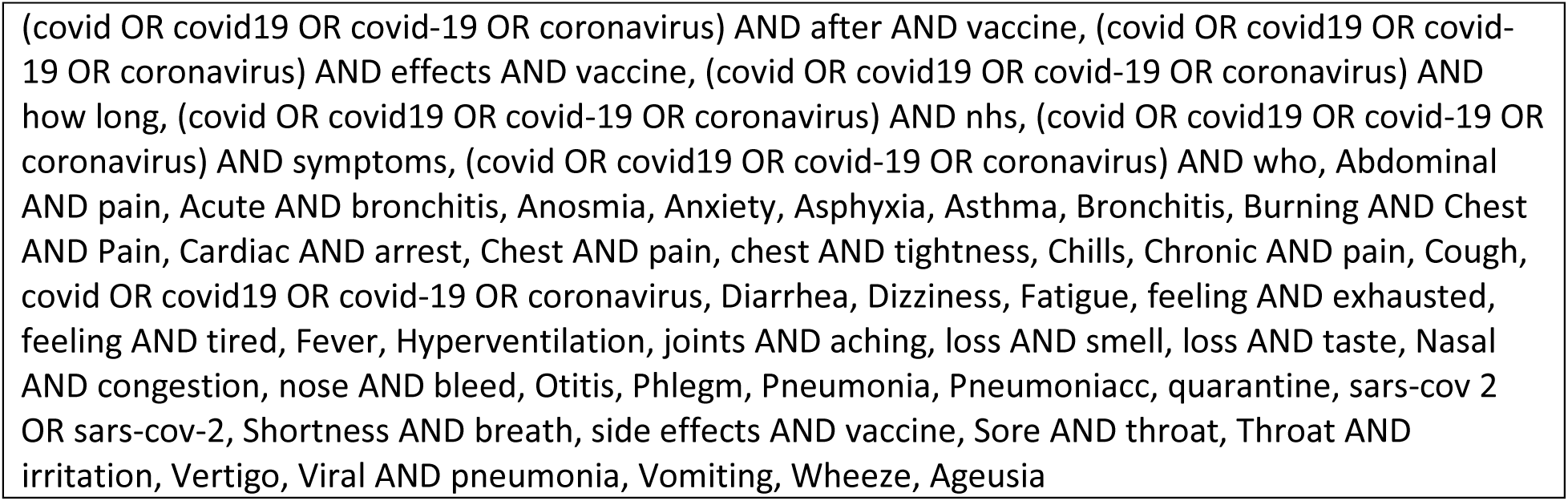
Search for the key list of Twitter.

**Table 3.**
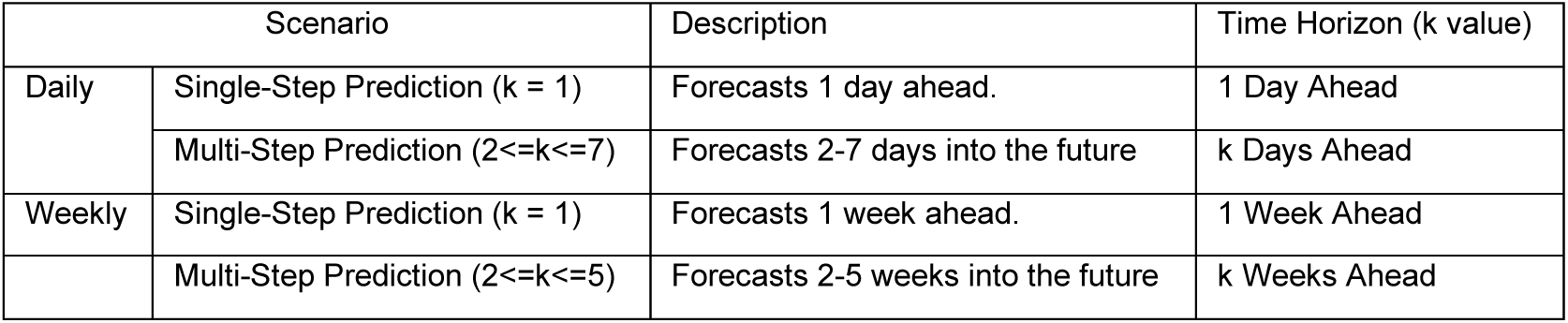
Summarizes the two prediction scenarios used in the training process.

Since Twitter’s geolocation metadata primarily provides city/town-level “place” identifiers rather than county or ZIP code information, we used Geocorr data [26], to map these locations to counties based on population distribution.

### 2.2 Methodology

The objective is to use historical data to train deep learning models. The models should identify the complex relationship between input features and future values of the target variable. The model will then forecast the target variable at future time steps. The functional relationship between the input features and the target variable is represented as:

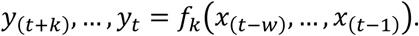

Where *y* represents the forecasted target variable (what we want to predict: cases or deaths), *k* denotes how far into the future the prediction is made (It is a hyperparameter determined during model training. For example, if *k*=7, the model predicts the next 7 days of data (e.g., daily case counts) based on the input sequence. For instance, one model might be trained to predict the next 7 days (*k*=7), while another might be trained to predict the next 14 days (*k*=14). This approach ensures that the model can adapt to different forecasting needs and time scales.), *x* represents the vector of observed input features (data used to make the prediction), the variable w signifies the look-back window size, meaning how much historical data the model uses for making the prediction. Also, fk represents the functional relationship between the input features (x) and the target variable (y) that the deep learning model aims to learn. This relationship is used to forecast the target variable at future time steps (k). In our framework, the input to the encoder is a sequence of length w, which corresponds to the look-back window. This sequence comprises the observed input features over the past w time steps, denoted as *x*_(*t*−*w*)_, …, *x*_(*t*−1)_. Each *x*_*t*_ is a feature vector with a dimensionality equal to the number of input features used in the model. These features, described in Section 2.1, include epidemiological indicators, demographic variables, and sentiment data derived from Twitter. Thus, the model ingests a matrix of shape (w × d), where d is the number of features, to learn patterns that inform the prediction of future target values. To adapt to different scenarios, our model’s input length can be adjusted based on the time series data. This flexibility is controlled by a hyperparameter that sets the maximum window size. This mathematical framework captures the core functionality of the model: identifying patterns and relationships in historical data to predict the target variable at future points.

We explored the methodology behind implementing deep learning models, particularly focusing on selecting neural network architectures like attention-based Transformers, which are adept at extracting both temporal and spatial relationships from the data. Figure 1 shows the schematic of our forecasting model. It consists of an input embedding and an encoder part. We adopted encoder-only architecture instead of the traditional encoder-decoder design. This choice simplifies the model, reduces training time, and improves performance by emphasizing direct representation learning over autoregressive decoding. Since our input data is structured, temporally aligned, and preprocessed, the decoder component was not essential and, in fact, introduced risks of overfitting and instability in multi-step forecasting. The encoder-only approach more effectively captures temporal patterns and cross-feature dependencies without the added complexity of sequence generation, leading to greater accuracy and robustness in predictions.

**Figure 1.**
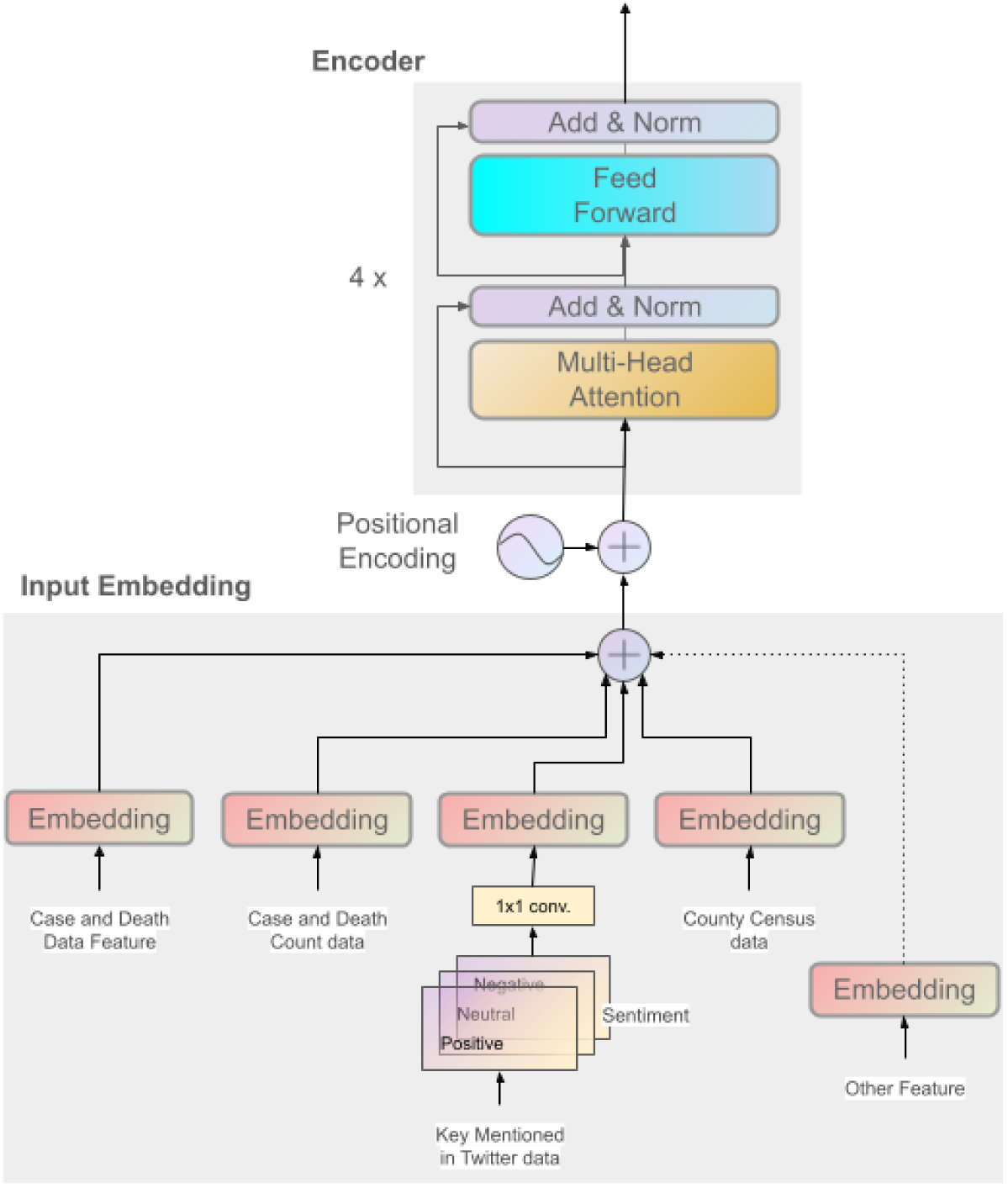
Architectural Configuration of the Forecasting Model Based on Transformer

#### 2.2.1 Self-attention mechanisms

This section focuses on how deep learning models, specifically attention-based Transformers, are implemented to extract temporal and spatial relationships from data. Transformers, known for their success in natural language processing, are used to capture long-term dependencies in time series data, outperforming traditional methods like RNNs and LSTMs in handling long sequences and training speed. [27].

Unlike the step-by-step approach of RNNs, Transformers utilize positional encoding. Positional encoding embeds the order of information into the data, allowing them to understand the time flow within the input. The encoder processes historical data (determined by window size ‘w’) and creates a feature vector, which the decoder uses for predictions. During training, the decoder receives the encoder’s output and the actual future data, enabling the model to learn from both past and future information. For a comprehensive understanding of the Transformer architecture and its attention mechanism, please refer to [27].

#### 2.2.2 Heterogeneous data integration, multi-source data fusion

For a more robust and precise forecasting model, we integrate data from diverse sources (e.g., social media sentiment) and formats (time series, text). Time series data reveals trends and patterns, while text data from social media and news provides context and sentiment, enriching the model’s perspective. For example, social media posts during a health crisis can offer early insights into public perception, potentially influencing the trajectory of the outbreak. Public discussions about symptoms, health risks, or healthcare access can signal emerging outbreaks or hotspots before official data confirms them. Analyzing sentiment related to illness, isolation, or healthcare-seeking behavior can provide valuable information about the disease’s geographic and temporal spread [28]. This comprehensive integration allows the model to discern complex relationships, capturing both quantitative and qualitative dimensions, and consequently, refining its predictive capabilities.

#### 2.2.3 Employing multi-level and multi-scale attention mechanisms in time series analysis

Transformer architecture processes input sequences over time using self-attention mechanisms, which allow the model to capture temporal dependencies and patterns in the data. While the transformer does not explicitly extract frequency-domain information (e.g., through spectral analysis), its attention mechanisms can implicitly learn to focus on periodic or trend-related behaviors in the input sequence. This implicit capability enables the model to handle time-series data effectively without requiring explicit modifications for frequency analysis.

#### 2.2.4 Combining traditional CNN methods with transformer technology

CNNs are used for preprocessing Twitter data for sentiment analysis (negative, positive, and neutral). This processed data is then fed into the Transformer model. The proposed model is a hybrid architecture that merges the feature extraction capabilities of CNNs with the powerful learning ability of Transformer networks.

## 3 Results

### 3.1 Model Training and Validation

The goal of our analysis is to predict current and future Covid-19 cases and deaths based on real-time data collected through X (formerly Twitter), historical Covid-19 cases and deaths, and community-level information (see **Section 2.1** for additional detail). To build a robust model, we addressed the inherent variability within our data. This variability stems from the inclusion of data from diverse sources with different formats and update frequencies (heterogeneity). Firstly, we acknowledged the inconsistent resolution (daily vs. weekly) across counties within the dataset. To address this, we applied a smoothing technique, specifically moving average, to ensure all data was represented at a daily resolution. Second, we addressed potential biases inherent in using diverse data sources. These biases can arise from inconsistencies in data formats, units, and time periods. Additionally, disparities in data collection across different regions and demographics can skew results. Extreme values, if left unmitigated, can disproportionately influence findings. Furthermore, changes in technology and other factors over time can render data collected at different periods incomparable. To mitigate this, we employed a random sampling approach. This approach involved randomly selecting a county’s data for inclusion in both the training and test sets. Additionally, we introduced randomness in the selection of start dates for these sets. This further enhances the generalizability of the model by ensuring it can learn from diverse data starting points. This randomization offers two key benefits. First, it injects temporal variability into the training process. By exposing the model to data from different starting points, we ensure it can capture the evolving dynamics of the pandemic, including any temporal trends or shifts in the data. Since training a model on data with a fixed start date could lead to the model memorizing specific patterns from that period rather than developing a broader understanding of pandemic dynamics. Second, given the limited dataset, employing random selection for training data helps prevent overfitting. This approach ensures the model encounters a diverse representation of the data during training, fostering better generalization to unseen examples.

We extend the randomization strategy to the data window length for training and testing. This exposes the model to various historical data durations, enhancing its adaptability to different timeframes. By encountering diverse temporal contexts, the model strengthens its ability to generalize and make accurate forecasts across a wider range of scenarios.

Our approach incorporates randomness when selecting data sources, start dates, and window lengths for training. This deliberate strategy injects variability into the training process. By encountering a wider range of data characteristics (county, date range, historical depth), the model is better equipped to handle the complexities of real-world data [29]. This randomization not only fosters a more versatile and adaptable model, but also strengthens its generalizability, allowing for accurate predictions across diverse scenarios. In essence, this approach enhances the model’s robustness by making it less susceptible to biases or specific data patterns.

#### 3.1.1 Architectural Details and Training Settings

The proposed model (Figure 1) is a custom architecture featuring input embeddings that project feature vectors into a 512-dimensional space. This is followed by 4-layer transformer encoders, each incorporating 8 attention heads, 64-dimensional queries, keys, and values, 1024-dimensional feedforward networks, and a dropout rate of 0.1. To incorporate temporal awareness, sinusoidal positional encodings are added to the input embeddings. The encoder generates predictions for the subsequent *k* time steps, which are then linearly projected to outputs. The Twitter data is processed into three sentiment channels—positive, negative, and neutral—forming a 2D matrix of shape (w×3), where *w* is the look-back window. A 2D CNN with a 1×1 kernel and ReLU activation projects inputs into a higher-dimensional space with a number of filters equal to the model’s embedding dimension (e.g., 512). The CNN output, a (*w* × *d*) matrix, is passed through a fully connected layer to align with the model’s embedding space. This sentiment representation is then added to the embeddings of other input features, enabling seamless integration of sentiment and traditional data into the model’s unified input. Training employs the Adam optimizer with a learning rate of 1e-4 and a weight decay of 1e-5, utilizing a warmup learning rate schedule and a batch size of 32. The model is trained for up to 200 epochs, with early stopping based on validation loss. Optimization is guided by the root mean squared error loss, and regularization is applied throughout the model via dropout and layer normalization.

#### 3.1.2 Training

To guarantee the reliability of our results, we strategically divided our dataset into two distinct periods. The first period, encompassing data from January 1, 2020, to November 30, 2021, served as the training set. This data allowed our model to learn historical trends and patterns, forming the foundation for its predictive capabilities. We evaluated our model’s performance using a validation set encompassing the period from December 1, 2021, to March 1, 2023. We divided this validation set into three sub-ranges corresponding to three SARS-CoV-2 variants: December 1, 2021, through March 31, 2022 (Omicron B.1.1.529 variant); May 1, 2022, through October 31, 2022 (Omicron BA.4 and BA.5 variants); and December 1, 2022, through February 28, 2023 (Omicron BQ.1 and XBB.1.5 variants). By testing the model’s predictions against this unseen data, we were able to assess its accuracy and effectiveness in forecasting Covid-19 trends for the specified timeframe. This two-part approach not only facilitated the fine-tuning of our model but also provided a robust evaluation of its real-world generalizability.

The transformers in our model are trained from scratch rather than being pretrained. Importantly, the training process is designed to handle data from all counties simultaneously, rather than training separate models for each county.

##### Look-Back Window Size (w)

The look-back window size (*w*) represents the length of historical data used as input for the forecasting. It determines how much past information the model considers when making predictions about future Covid-19 cases. The look-back window allows the model to identify trends and patterns in historical case data. A larger window provides more context, potentially enabling the model to recognize more complex patterns and make more informed predictions. While a larger window might seem advantageous, it can also introduce noise and make the model less sensitive to recent changes. A smaller window might be more responsive to short-term fluctuations but may miss longer-term trends. The model’s maximum look-back window is a hyperparameter, meaning it can be adjusted during the training process. This allows us to experiment with different window sizes and find the optimal balance for a specific prediction task. To capture historical trends while training the model, we employ a maximum lookback window of *w = 16* days (or weeks, depending on the model’s daily or weekly resolution). This window defines the maximum historical data span the model considers during training. While the actual training or prediction process might utilize all available data within this window, it won’t exceed this limit. In our experiments, the maximum look-back window was set to 16 days for daily prediction and 5 weeks for weekly prediction.

##### Single-Step vs. Multi-Step Predictions

Our training process involves two prediction scenarios: single-step and multi-step. Single-step predictions (denoted by *k = 1*) aim to forecast the value at the next step. In multi-step prediction, the model predicts a sequence of future values (e.g., the next *k* days or weeks) in a single forward pass. For multi-step predictions, we trained separate models for different prediction horizons (*k*), allowing each model to specialize in forecasting a specific time range. This approach avoids the error accumulation that can occur in autoregressive models and ensures that the model captures dependencies across the entire prediction horizon. The maximum multi-step predictions in our experiments aimed to forecast results 7 days and 5 weeks ahead.

### 3.2 Predict multiple timesteps ahead

We analyze the model’s accuracy in predicting peak timing compared to actual occurrences across three distinct periods: December 1, 2021 – March 31, 2022, May 1, 2022 – October 31, 2022, and December 1, 2022 – February 28, 2023, using three history data: November 1, 2021 – March 31, 2022, April 1, 2022 – October 30, 2022, and November 1, 2022 – February 28, 2023. We investigated how the model’s performance changes as we predict further into the future (*k*). Figure 2 and Figure 3 present the comparison of total daily and weekly actual cases and deaths for three-time frames for all counties. As expected, the model’s accuracy generally decreases as *k* increases. The figures specifically highlight the model’s ability to forecast trends over different timeframes. The visual representation in the figures allows for a direct assessment of the model’s accuracy in tracking the pandemic’s trajectory, showing how closely the predicted lines align with the actual observed data.

**Figure 2.**
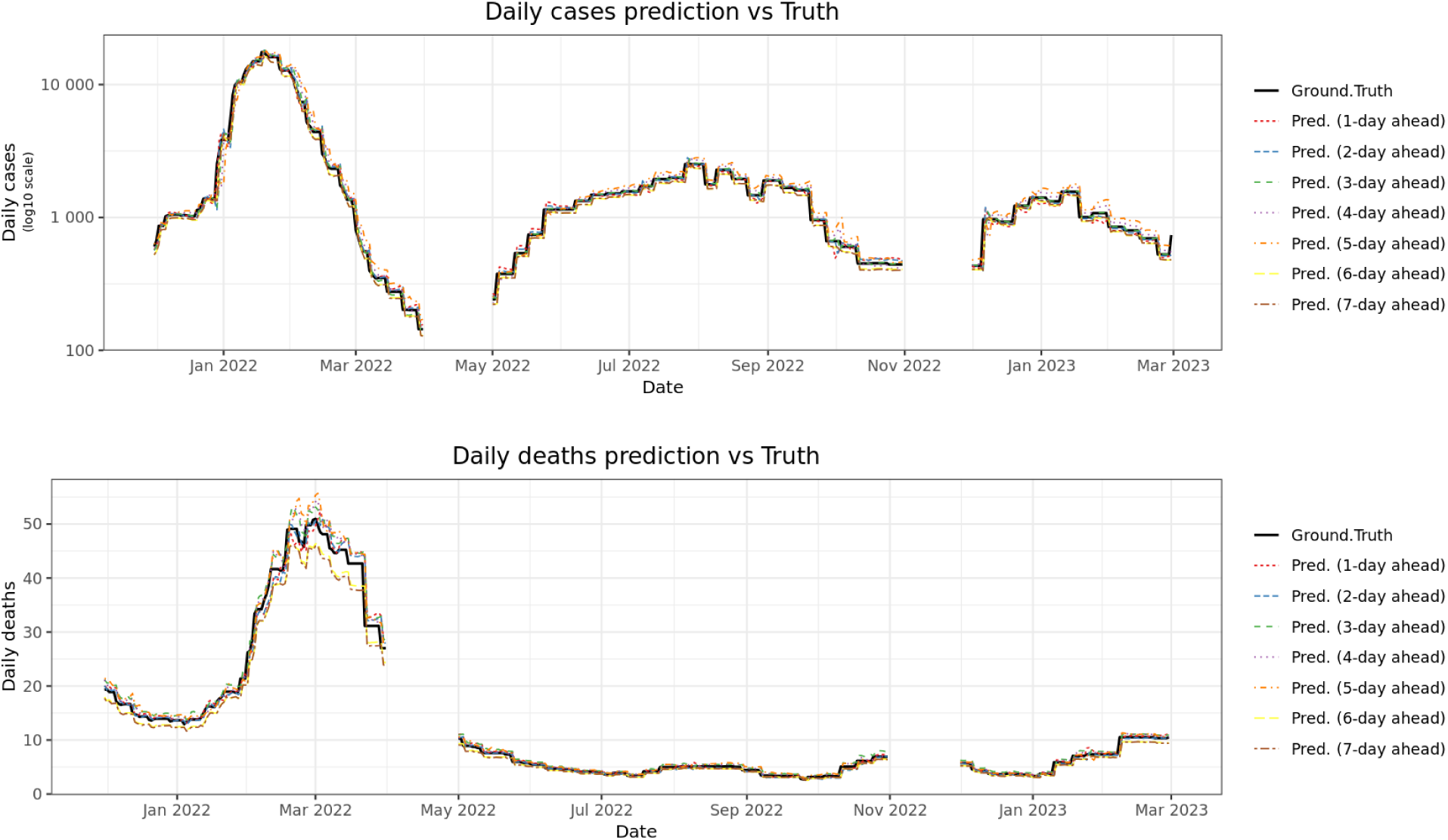
Comparison of total daily predicted and actual values of Covid-19 positive cases and deaths. Top: daily case prediction, Bottom: daily death prediction.

**Figure 3.**
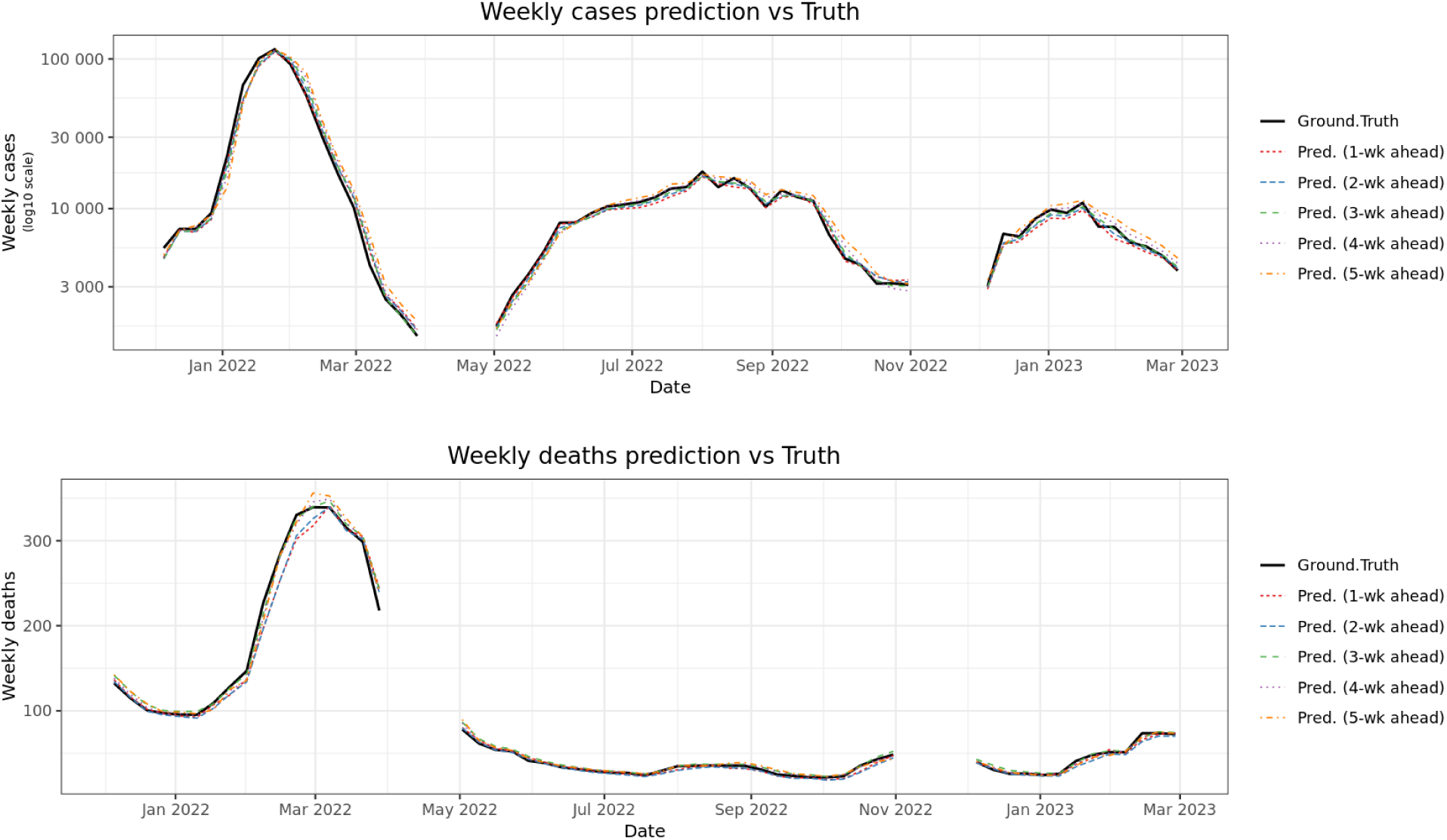
Comparison of total weekly predicted and actual values of Covid-19 positive cases and deaths. Top: weekly case prediction, Bottom: weekly death prediction.

Average prediction accuracy across all counties is presented in Figure 2. We consistently observe higher accuracy with lower *k* values. The predictions closely follow observed case trends and death trends, except for overprediction of deaths at the tail of the first Omicron wave (March 2022). Performance accuracy decreases as *k* increases for weekly predictions (Figure 3).

### 3.3 Performance Metrics

#### 3.3.1 Performance Metrics (PA)

We evaluated the agreement between predicted and observed cases or deaths using the following percentage agreement metric. These metrics provide insight into the temporal reliability of the forecasting model, with higher PA values indicating stronger predictive alignment. This evaluation is critical for assessing the model’s performance in short-term epidemic forecasting and informing public health decision-making at the local level. The PA formula is

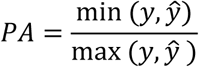

*ŷ* represents predicted value, *y* represent observed value.

The PA metric of a county is the average of the agreement over the predicted date range.

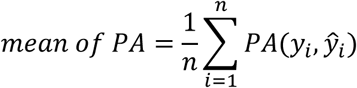

*ŷ*_*i*_ represents predicted value, *y*_*i*_ represent observed value, *n* represents total number of observations (the total days or weeks of the predicted date range in each county).

Table 4 reports the county-level Percentage Agreement (PA) metrics for COVID-19 case and death predictions across a seven-day forecast horizon, with each column representing the PA for predictions made one to seven days ahead. The median PA for daily case predictions generally shows a decreasing trend as the prediction horizon extends from Day 1 to Day 7 across all the observed time periods. This decline suggests that the accuracy of predicting daily case numbers tends to diminish as the forecast extends further into the future, which is a common characteristic of forecasting models due to increasing uncertainty over longer periods. Comparing the median PA values across different time periods for each prediction day reveals that the accuracy appears to be generally higher in the initial phases of the study compared to the later periods. This could indicate a change in the predictability of case numbers as the pandemic evolved, possibly due to factors like the emergence of new variants or shifts in public health interventions. The “deaths” section of Table 4 provides similar data for daily death predictions across the same time periods and prediction horizons. The trend in median Percentage Agreement for daily death predictions is less consistently decreasing with the prediction horizon compared to cases. This non-linear trend might reflect the delayed impact of case surges on mortality figures. Comparing across time periods, the median PA for death predictions is generally high in the earlier periods (12/01/2021 and 03/31/2022), often exceeding 80% even for longer prediction horizons. However, there is a noticeable decline in accuracy in the later periods. This decrease could be attributed to various factors, such as increased vaccination rates altering the case-fatality ratio or changes in reporting practices for deaths.

**Table 4.**
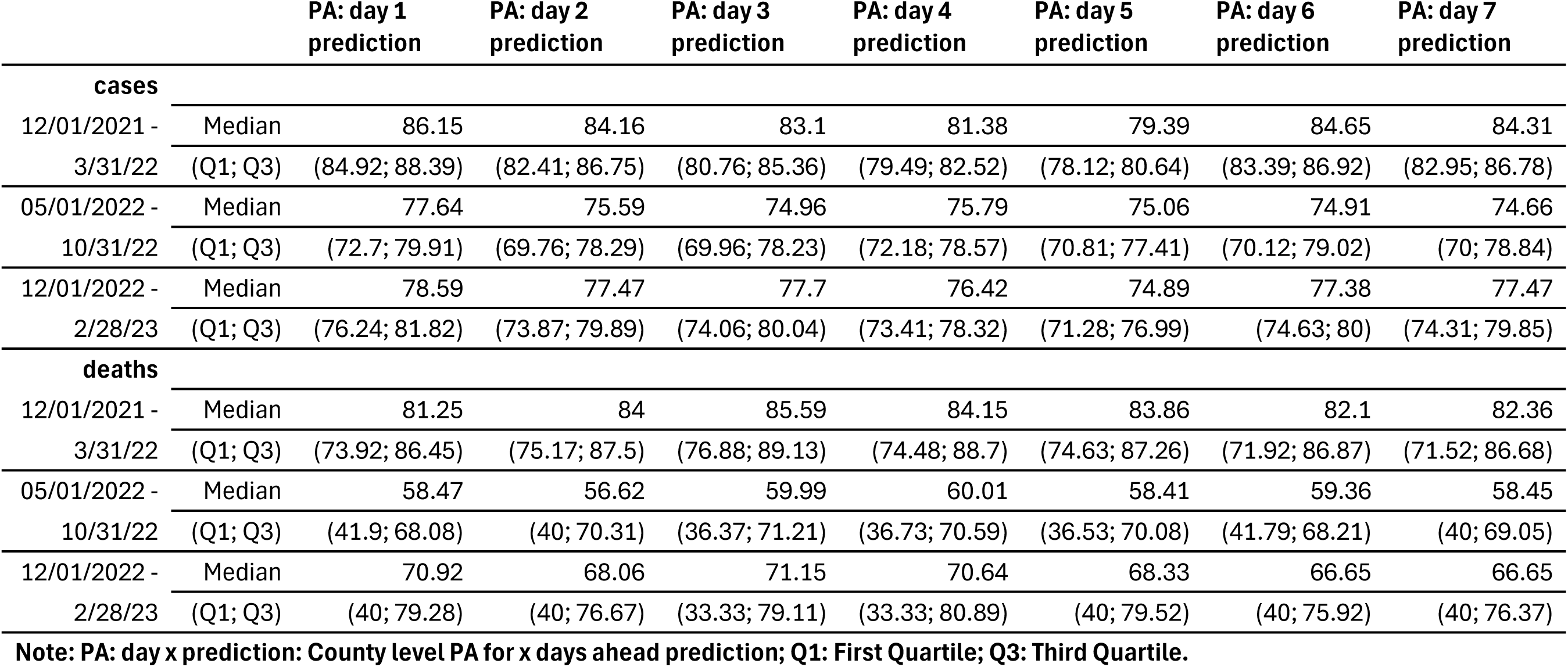
County-Level Percentage Agreement (PA) and interquartile range (IQR) for daily prediction.

|  |  | PA: day 1<br>prediction | PA: day 2<br>prediction | PA: day 3<br>prediction | PA: day 4<br>prediction | PA: day 5<br>prediction | PA: day 6<br>prediction | PA: day 7<br>prediction |
| --- | --- | --- | --- | --- | --- | --- | --- | --- |
| **cases** |  |  |  |  |  |  |  |  |
| 12/01/2021 - | Median | 86.15 | 84.16 | 83.1 | 81.38 | 79.39 | 84.65 | 84.31 |
| 3/31/22 | (Q1; Q3) | (84.92; 88.39) | (82.41; 86.75) | (80.76; 85.36) | (79.49; 82.52) | (78.12; 80.64) | (83.39; 86.92) | (82.95; 86.78) |
| 05/01/2022 - | Median | 77.64 | 75.59 | 74.96 | 75.79 | 75.06 | 74.91 | 74.66 |
| 10/31/22 | (Q1; Q3) | (72.7; 79.91) | (69.76; 78.29) | (69.96; 78.23) | (72.18; 78.57) | (70.81; 77.41) | (70.12; 79.02) | (70; 78.84) |
| 12/01/2022 - | Median | 78.59 | 77.47 | 77.7 | 76.42 | 74.89 | 77.38 | 77.47 |
| 2/28/23 | (Q1; Q3) | (76.24; 81.82) | (73.87; 79.89) | (74.06; 80.04) | (73.41; 78.32) | (71.28; 76.99) | (74.63; 80) | (74.31; 79.85) |
| **deaths** |  |  |  |  |  |  |  |  |
| 12/01/2021 - | Median | 81.25 | 84 | 85.59 | 84.15 | 83.86 | 82.1 | 82.36 |
| 3/31/22 | (Q1; Q3) | (73.92; 86.45) | (75.17; 87.5) | (76.88; 89.13) | (74.48; 88.7) | (74.63; 87.26) | (71.92; 86.87) | (71.52; 86.68) |
| 05/01/2022 - | Median | 58.47 | 56.62 | 59.99 | 60.01 | 58.41 | 59.36 | 58.45 |
| 10/31/22 | (Q1; Q3) | (41.9; 68.08) | (40; 70.31) | (36.37; 71.21) | (36.73; 70.59) | (36.53; 70.08) | (41.79; 68.21) | (40; 69.05) |
| 12/01/2022 - | Median | 70.92 | 68.06 | 71.15 | 70.64 | 68.33 | 66.65 | 66.65 |
| 2/28/23 | (Q1; Q3) | (40; 79.28) | (40; 76.67) | (33.33; 79.11) | (33.33; 80.89) | (40; 79.52) | (40; 75.92) | (40; 76.37) |

**Note: PA: day x prediction: County level PA for x days ahead prediction; Q1: First Quartile; Q3: Third Quartile.**

Similarly, Table 5 provides the corresponding PA metrics for forecasts made one to five weeks ahead, offering a parallel evaluation of model performance over longer-term prediction windows. Results are shown for three different evaluation periods, highlighting temporal changes in model performance. In general, case predictions exhibit higher PA than death predictions, particularly during the earlier periods of evaluation. The decreasing PA values over longer forecast horizons reflect the expected increase in uncertainty with longer-term predictions. The reported IQRs provide additional information about the variability across counties, emphasizing regions where forecasts were reliable. Weekly forecasts of deaths tend to achieve higher PA values than those of cases, reflecting greater stability in death predictions over longer timescales. As the prediction horizon extends from one to five weeks, a gradual decline in PA is observed, consistent with the increasing uncertainty inherent in longer-term forecasts. The overall accuracy for weekly case predictions appears to be reasonably high across the study period, and the consistency across counties, as reflected by the IQR, is generally better than that of daily case predictions. The accuracy remains reasonably good, suggesting that weekly aggregation might provide a more stable level of accuracy over longer forecast horizons compared to daily predictions. The median PA for weekly death predictions remains high across all time periods and prediction horizons compared to daily death predictions.

**Table 5.** County-Level Percentage Agreement (PA) and interquartile range (IQR) for weekly prediction.

|  |  | PA: week 1<br>prediction | PA: week 2<br>prediction | PA: week 3<br>prediction | PA: week 4<br>prediction | PA: week 5<br>prediction |
| --- | --- | --- | --- | --- | --- | --- |
| **cases** |  |  |  |  |  |  |
| 12/01/2021 - | Median | 73.98 | 73.34 | 72.48 | 70.32 | 68.72 |
| 3/31/22 | (Q1; Q3) | (71.36; 75.4) | (71.32; 75.31) | (69.64; 73.96) | (68.96; 72.48) | (66.66; 71) |
| 05/01/2022 - | Median | 82.59 | 82.15 | 81.2 | 80.95 | 79.6 |
| 10/31/22 | (Q1; Q3) | (78.77; 85.13) | (78.1; 84.19) | (76.66; 83.3) | (76.98; 82.42) | (75.48; 81.73) |
| 12/01/2022 - | Median | 81.62 | 82.01 | 81.69 | 80.34 | 78.34 |
| 2/28/23 | (Q1; Q3) | (78.85; 83.82) | (79.86; 84.48) | (79.67; 84.56) | (78.2; 82.66) | (74.66; 79.79) |
| **deaths** |  |  |  |  |  |  |
| 12/01/2021 - | Median | 84.96 | 86.42 | 87.83 | 87.66 | 86.84 |
| 3/31/22 | (Q1; Q3) | (81.49; 87.55) | (83.11; 88.1) | (84.78; 89.4) | (84.65; 89.78) | (84.17; 88.6) |
| 05/01/2022 - | Median | 83.2 | 82.21 | 85.46 | 86.51 | 84.35 |
| 10/31/22 | (Q1; Q3) | (80.71; 85.65) | (79.42; 84.64) | (83.96; 87.02) | (84.64; 88.41) | (82.78; 87.07) |
| 12/01/2022 - | Median | 86.34 | 84.54 | 86.52 | 88.02 | 87.23 |
| 2/28/23 | (Q1; Q3) | (83.25; 89.36) | (80.55; 88.65) | (83.33; 89.36) | (85.01; 91.42) | (83.58; 90.01) |

**Note: PA: week x prediction: County level PA for x weeks ahead prediction; Q1: First Quartile; Q3: Third Quartile.**

#### 3.3.2 Root Mean Squared Error (RMSE) and Mean Absolute Error (MAE)

The Root Mean Square Error metrics helps assess how prediction error evolves over time.

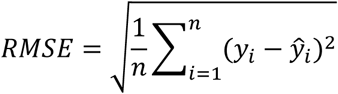

RMSE is a commonly used metric to evaluate model prediction accuracy. A lower RMSE indicates better performance.

Table A1 reports the county-level RMSE and interquartile range (IQR) for daily COVID-19 case and death predictions over a seven-day forecast horizon. The results show that RMSEs for case predictions tend to increase slightly with longer forecast horizons, reflecting the accumulation of prediction uncertainty over time. Table A2 presents the county-level RMSE and IQR for weekly COVID-19 case and death forecasts, extending from one-week to five-week ahead predictions. As expected, RMSE values for case predictions increase with longer forecast windows, indicating greater difficulty in accurately predicting further into the future.

We also employ Mean Absolute Error (MAE) metric. MAE measures the average absolute deviation between predicted and observed values:

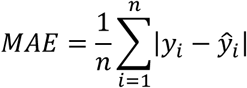

Table A3 and Table A4 summarize the county-level MAE and IQR for daily and weekly predictions of COVID-19 cases and deaths. For daily case predictions the median MAE was relatively high between December 2021 and March 2022 (ranging from 4.04 to 7.5 cases), but steadily declined over time, reaching approximately 1 to 2 cases by late 2022 and early 2023. As expected, prediction errors increased slightly with longer forecast horizons (e.g., day 7 predictions were generally less accurate than day 1 predictions). Similarly, weekly case predictions showed initially large errors during the early period (median MAEs between 91.57 and 111.94 cases) but decreased substantially over time, with medians dropping to approximately 15–20 cases per week by early 2023.

### 3.4 Impact of incorporating twitter (x) data input and Comparison with the Persistence Model

To assess the impact of incorporating specific input data into our forecasting model, we performed comparative experiments across three model configurations. First, we evaluated our full model, which includes the Twitter (X) data input and compared it to a similar model omitting Twitter (X) data. This model was trained with the same model architecture used in the full model but excluded the Twitter(X) data input to isolate its effect. Then, we implemented a baseline persistence model, which predicts future values by simply repeating the most recent observation. The persistence model serves as a standard benchmark in time series forecasting, particularly in epidemiological contexts, due to its simplicity and effectiveness in stable or slowly evolving systems. By directly forwarding the current value as the prediction for the next time step, it avoids any modeling of temporal dynamics. Through this comparison, we aim to quantify both the improvement offered by our model over a naive baseline and the specific contribution of the X data input in enhancing predictive accuracy. Comprehensive performance metrics, including Percentage Agreement (PA), Root Mean Square Error (RMSE), and Mean Absolute Error (MAE), for both the persistence model and the full model without the Twitter(X) input are presented in Appendix Tables A5–A16.

Figures 4 and 5 show the percentage agreement of daily and weekly Covid-19 case and death predictions with and without social media data and the persistence model across three time periods. During the first time period (omicron wave 1), incorporating social media data improved median prediction accuracy by 4.94% to 10.50% (cases) and −1.04% to 4.70% (deaths) for predictions of 1 to 7 days ahead. Compared with the Persistence model, our model improved median prediction accuracy by 0.80% to 39.93% (cases) and −4.39% to 10.30% (deaths). In the second time period (omicron wave 2), incorporation of social media data yielded improvement ranging from --4.22% to 4.06% (cases) and - 2.75% to 4.27% (deaths). Compared with the Persistence model, our model improved median prediction accuracy by −1.42% to 2.25% (cases) and −4.71% to 3.47% (deaths). In the third time period (omicron wave 3), incorporating social media data yielded improvement ranging from −3.57% to 3.53% (cases) and 0.50% to 8.20% (deaths). Compared with the Persistence model, our model improved median prediction accuracy by −1.76% to 6.74.% (cases) and −2.07% to 4.54% (deaths). For 7 days peak prediction, improvement ranged from 1.19% to 9.34% (cases) and 3.77% to 4.89% (deaths) across all three waves.

**Figure 4.**
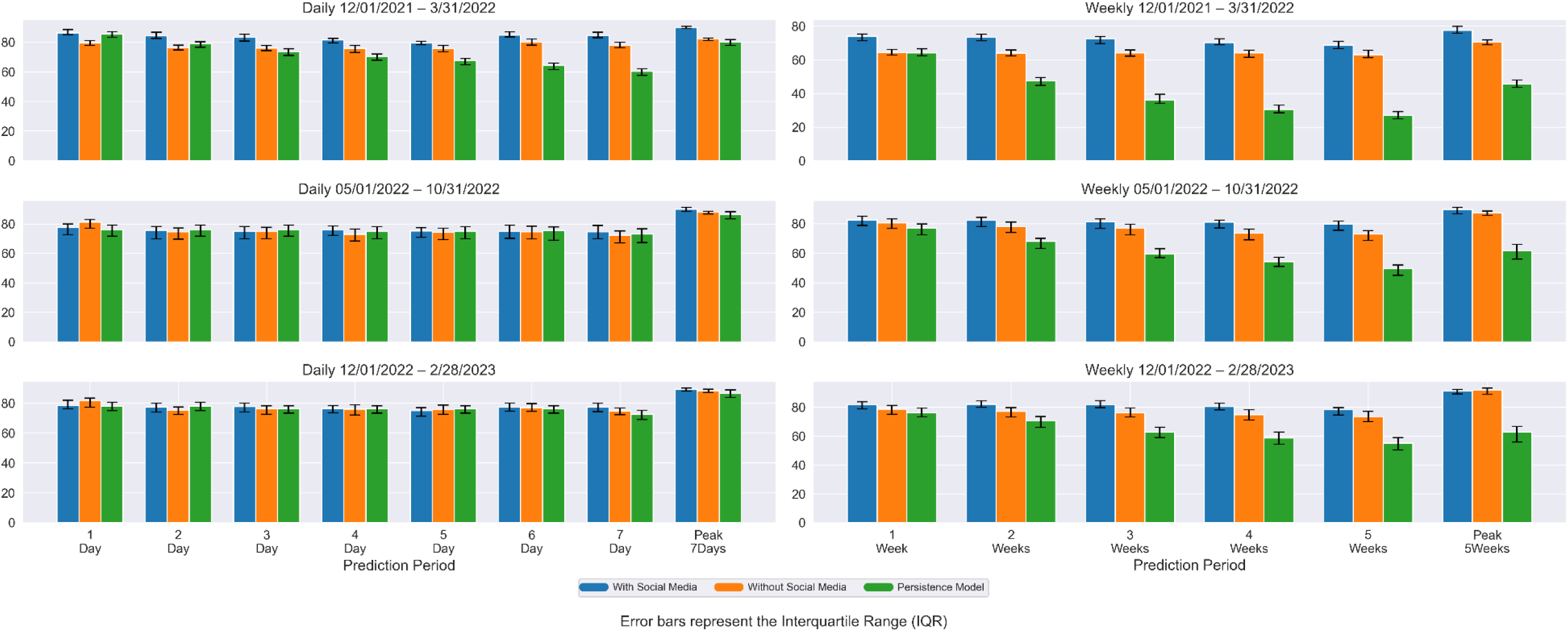
Median percentage agreement (PA) of daily and weekly Covid-19 case predictions with and without social media data across three time periods. The left three sub-graphs compare predictions of 1 to 7 days ahead and 7-days peak, right 3 sub-graphs compare predictions of 1 to 5 weeks ahead and 5-weeks peak. The error bar represents the interquartile range for PA across counties.

**Figure 5.**
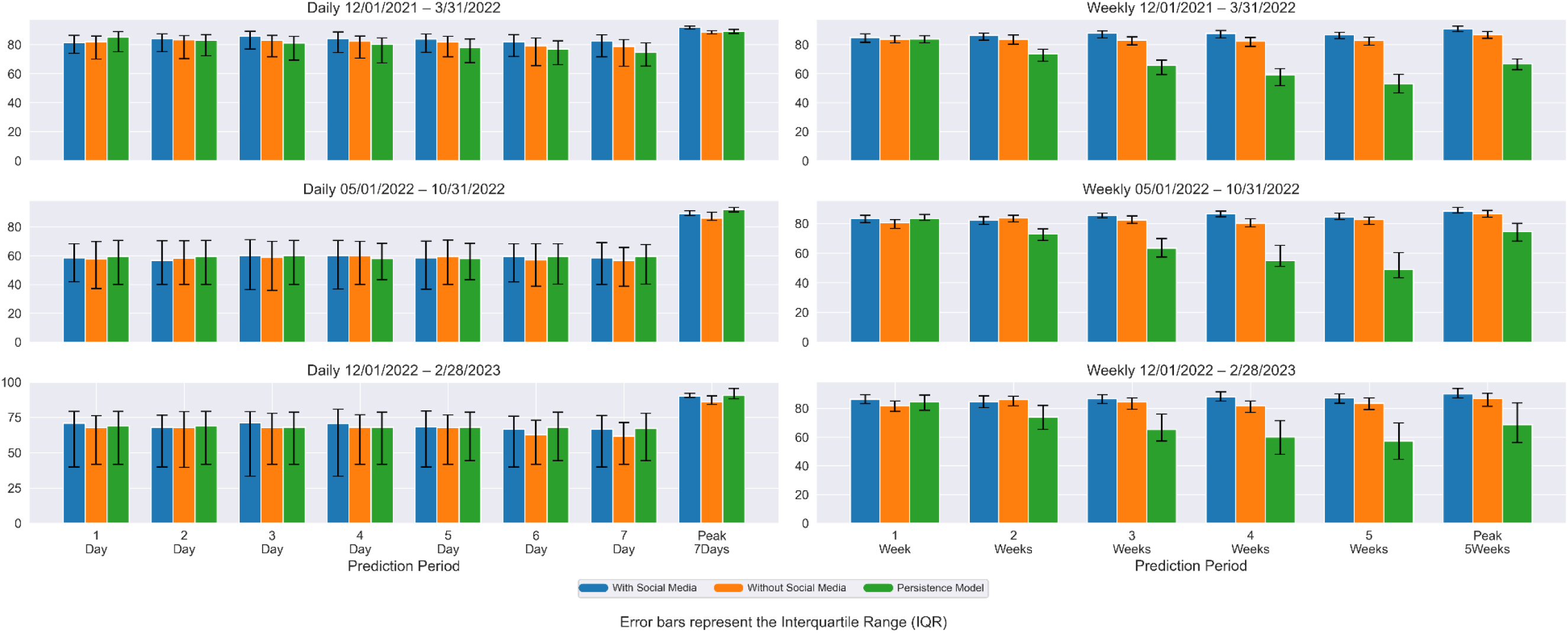
Median percentage agreement (PA) of daily and weekly Covid-19 death predictions with and without social media data across three time periods. The left three sub-graphs compare predictions of 1 to 7 days ahead and 7-days peak, right 3 sub-graphs compare predictions of 1 to 5 weeks ahead and 5-weeks peak. The error bar represents the interquartile range for PA across counties.

For weekly prediction, during the first time period (omicron wave 1), incorporating social media data improved median prediction accuracy by 9.04% to 14.82% (cases) and 1.65% to6.62% (deaths) for predictions of 1 to 5 weeks ahead. Compared with the Persistence model, our model improved median prediction accuracy by 15.31% to 155.94% (cases) and 1.36% to 63.85% (deaths). In the second time period (omicron wave 2), incorporating social media data yielded improvement ranging from −2.69% to 10.09% (cases) and −1.83% to 7.94% (deaths). Compared with the Persistence model, our model improved median prediction accuracy by 7.15% to 60.97% (cases) and −0.12% to 73.13% (deaths). In the third time period (omicron wave 3), incorporating social media data yielded improvement ranging from - 0.49% to 7.52% (cases) and --1.62% to 7.66% (deaths). Compared with the Persistence model, our model improved median prediction accuracy by 7.28% to 45.27% (cases) and 2.27% to 52.63% (deaths). For 5 weeks peak prediction, improvement ranged from −0.49% to 9.81% (cases) and 2.08% to 4.54% (deaths) across all three waves.

Performance comparisons across additional metrics (RMSE, MAE) are provided in Supplementary Tables A1–A4 and A11–A16. Across all forecasting horizons and evaluation metrics, our model consistently outperformed the persistence model. For longer-term forecasts, where persistence models typically exhibit degraded accuracy, our model achieved substantial improvements: a 34% increase in PA (81.9% vs. 61.2% average), a 64% reduction in RMSE (35.1 vs. 98.4 average), and a 68% reduction in MAE (23.0 vs. 72.1 average). These gains were most notable during epidemiological transitions (e.g., onset and decline phases), where the incorporation of leading indicators such as sentiment trends enabled our model to anticipate trend shifts more effectively than the static persistence baseline. Even in short-term (7-day) forecasts, our approach yielded meaningful improvements, including a 4% improvement in PA accuracy (74.5% vs 71.6% average), a 46% reduction in RMSE (2.6 vs 4.8 average), and a 51% reduction in MAE (1.4 vs 2.8 average), while remaining suitable for real-time deployment.

Overall, incorporating county-level social media data significantly enhanced the prediction accuracy of both daily and weekly COVID-19 cases and deaths across various forecast horizons and pandemic waves. The improvements were most pronounced during the first Omicron wave, where the inclusion of social media led to consistently higher median accuracy compared to both models without social media and a persistence baseline. This suggests that social media signals were especially valuable during periods of rapid epidemiological changes and heightened public discourse. While the magnitude of improvement diminished somewhat during the second and third Omicron waves—predictions generally remained more accurate with social media input. Notably, even modest gains in forecast accuracy, particularly for weekly and peak predictions, can support better-informed public health responses and resource allocation.

Figure 6 and Figure 7 present the predicted weekly peak cases and deaths for the period December 2021 to March 2023 using 1 week ahead prediction. Notably, these figures also depict the volume of Twitter mentions corresponding to the dates with the predicted peaks. This combined visualization allows us to investigate potential correlations between social media activity and forecasted peaks in cases and deaths, enabling us to leverage social media data to improve the accuracy of future infectious disease forecasts. In Figures 6 and 7, percentage agreement was computed based on comparing the predicted and observed cases and deaths during the week where the predicted peak occurred across the entire study period (December 2021 through February 2023). The Twitter mentions correlate with higher case and deaths in populous counties. The high agreement across most counties suggests that the predictive model is generally reliable, but areas with moderate agreement indicate room for model refinement.

**Figure 6.**
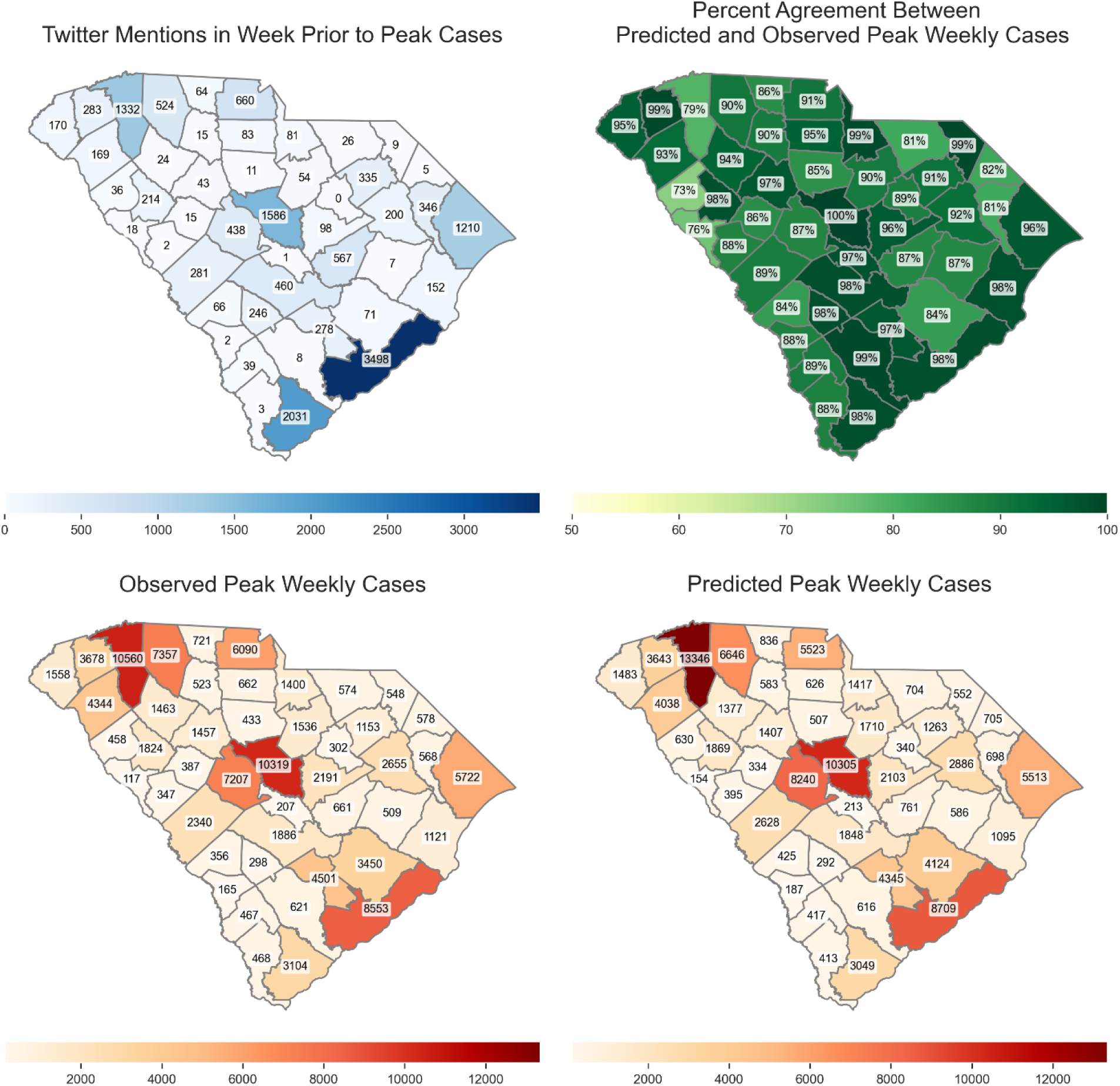
Peak cases weekly predicted, truth cases and twitter mention. The color scheme shows the predicted counts in each county.

**Figure 7.**
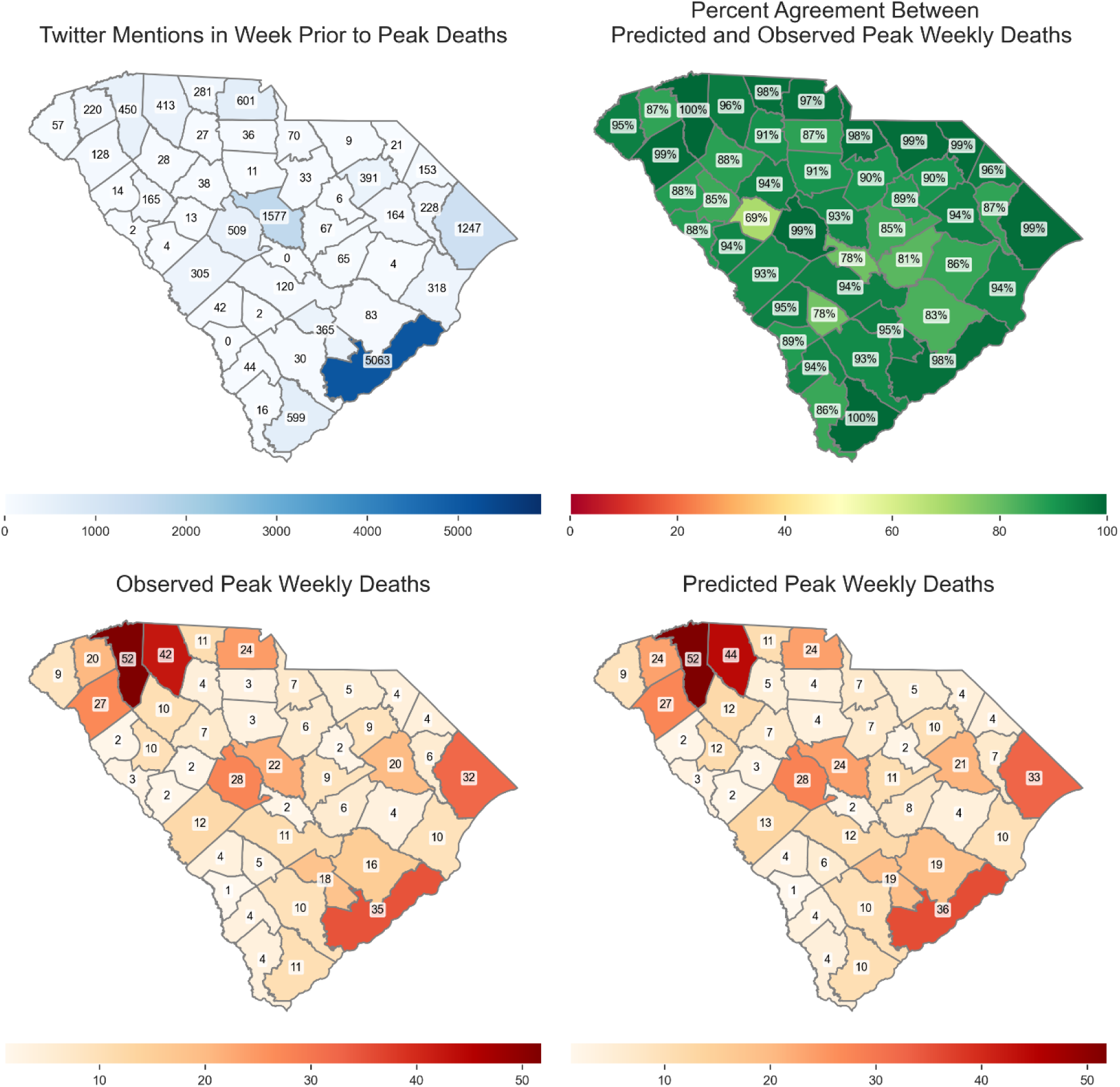
Peak deaths weekly predicted, truth cases and twitter mention. The color scheme shows the predicted counts in each county.

## 4 Discussion

The results of our research demonstrate the effectiveness of utilizing deep learning models for predicting trends in the Covid-19 pandemic. The Transformer architecture employed in our study exhibited strong performance in capturing both short- and long-term patterns within the data.

Moreover, our multi-source data fusion approach and multi-level and multi-scale attention mechanisms further enhanced the model’s accuracy and generalizability. The findings suggest that deep learning models can be integrated into public health surveillance systems to complement traditional epidemiological data and improve pandemic response efforts.

Case and death forecasting was highly accurate in most settings, with diminished model performance as the prediction window increased. The only exception is case prediction during the first Omicron wave, which resulted in substantially lower prediction accuracy. It is notable that, social media data particularly improves the accuracy of peak predictions, which is crucial for planning and response efforts. The comparison demonstrates the substantial advantages of integrating social media information into epidemiological models, enhancing both the precision and reliability of Covid-19 case predictions. This approach can be useful in better preparing for and responding to future public health challenges.

In considering the generalizability of our approach, it is important to note that although the model was trained using data from South Carolina, its architecture was intentionally designed to support adaptation to other geographic regions. The inclusion of demographic features such as population density, age distribution, and income level help account for regional variability, enhancing the model’s potential for broader application. Furthermore, input variables are processed using standardized methods at the national level, promoting consistency across different locations. To extend the model to other states or regions, retraining can be performed using locally sourced data while maintaining the same model structure. However, as demographic and epidemiological dynamics vary by location, model performance may differ accordingly. We therefore recommend localized retraining to ensure optimal predictive accuracy in new settings.

While national initiatives like the COVID-19 Forecasting Center [30] provide robust ensemble forecasts at the state and national levels, our research addresses an unmet need for localized forecasting by developing a county-level model for South Carolina. The model contributes three key elements: (1) fine-grained spatial resolution to support targeted local interventions; (2) integration of non-traditional data sources, including sentiment and demographic indicators from Twitter; and (3) significant performance improvements, up to 4% improvement in short-term (1-7 days) forecast accuracy and 34% improvement in long-term (1-5 weeks) forecast accuracy over the persistence model. While our current framework does not include hospitalization forecasts, it complements existing ensemble systems by providing high-resolution insights that are critical for micro-level decision making. Future work will explore hybrid approaches that combine the localized strengths of our model with the robustness of broader ensemble approaches to further enrich the pandemic forecasting ecosystem.

It is important to acknowledge the limitations in predicting the precise timing of peaks, especially for forecasts extending further into the future. The accuracy of the model’s predictions generally diminishes as the prediction window (*k*) increases. This trend is particularly noticeable when predicting the timing of peak cases and deaths. Furthermore, analysis consistently reveals a lag in the model’s peak predictions compared to the actual occurrences. This lag is evident across the three distinct periods examined. This recurring lag underscores the challenges in achieving perfect real-time alignment between predictions and the dynamic nature of pandemic outbreaks, even with sophisticated deep learning models. Several factors might contribute to these limitations. The dynamic and multifaceted nature of pandemic spread, influenced by evolving factors like viral mutations, public health interventions, and individual behavior, poses inherent challenges to predictive modeling. Additionally, the reliance on historical data, while crucial for training deep learning models, also implies a dependence on past trends that might not fully represent future pandemic behavior.

### 4.1 Limitations and Future Extensions

This study was retrospective and did not involve real-time deployment of the model. Since the New York Times stopped reporting COVID-19 data in March 2023 [31], live validation using the same data sources was not feasible. As a result, while the model performed well historically, its real-time accuracy remains untested. Additionally, while Twitter/X data provided marginal gains, its utility is context-dependent and requires continuous validation given platform dynamics, external changes in data sources—such as Twitter/X platform updates or shifts in search engine behavior—may weaken the relationship between input signals and outcomes over time.

It is also important to consider that social media activity can be influenced by a variety of factors besides the actual disease spread. News articles, for instance, can trigger spikes in social media conversations that might not directly reflect the number of cases. Additionally, public health interventions or changes in testing practices could also lead to fluctuations in reported cases that might not be perfectly captured by social media trends. To determine a more definitive relationship between social media mentions and predicted disease peaks, a more in-depth analysis would be required. This could involve: Investigating the time lag between social media mentions and subsequent case/death peaks could be informative.

Social media activity might precede or follow actual case increases, and understanding this time lag could be valuable for forecasting purposes. Analyzing how other factors (e.g., news reports, government policies, school closures) coincide with social media activity and case/death peaks. This broader analysis would provide a more comprehensive picture of the potential influences on disease spread and public discussion.

These shifts highlight the risk of performance degradation if models are not periodically retrained. However, this concern is not unique to social media data; it applies to any evolving data source. For instance, algorithmic changes in Google Trends or shifts in clinical reporting practices—such as changes in how or when cases are recorded—can similarly disrupt learned patterns and reduce forecasting accuracy over time. While our current study focused on evaluating generalizability without retraining, future work should explore the benefits of regular model updates. As pandemic dynamics shift with new variants, public health responses, and behavioral changes, retraining the model at set intervals or through online learning could improve its adaptability and maintain high forecasting accuracy. To improve adaptability, models should incorporate real-time data and support routine retraining to reflect changes in behavior, platforms, and virus dynamics. Expanding data sources—such as incorporation of electronic health records (EHR) or wastewater surveillance—could further strengthen predictive accuracy and generalizability in future outbreaks.

For example, with recent CDC policy changes no longer requiring hospitals to publish COVID-19 hospitalization data, or the NYT no longer reporting daily cases and deaths, are current modeling platform does not have the input features it needs for real-time prediction. Recent work has demonstrated the potential of using real-time EHR for nowcasting statewide hospitalizations from local health system data [32]. Our proposed framework can be adapted to this setting for infectious disease forecasting.

One of the most significant and addressable limitations of this study is the inconsistent temporal and spatial resolution across data sources. For example, epidemiologic data were reported daily, while some community-level contextual factors and social media indicators had irregular update intervals or were aggregated differently. Although smoothing and temporal alignment techniques were applied, mismatches in granularity may have caused the model to overlook short-term surges or fail to distinguish local anomalies. This limitation could be mitigated in future work by prioritizing data sources with consistent temporal frequency or by applying attention mechanisms that explicitly account for temporal gaps or spatial heterogeneity in the input sequences.

Finally, while our model achieves strong performance in COVID-19 prediction, it still has some limitations that deserve attention. The use of global self-attention mechanism enables comprehensive pattern recognition of all input features and improves computational efficiency by avoiding parameter-heavy channel-wise attention mechanism. However, this architecture may miss subtle but important feature interactions (e.g., the interaction between sentiment changes and case, death trends) and has limited interpretability compared to explicit channel-wise mechanisms. In addition, Transformer requires a large dataset to effectively learn such interactions. To address these challenges, future work may combine hybrid attention mechanisms, interpretable modules, and sparse feature-aware attention patterns to enhance learning ability and interpretability.

### 4.2 Conclusion

The ability to conduct accurate real-time forecasting at granular geographic regions is often hampered by both methodology and lack of relevant data for medically underserved areas. Due to their ability to identify complex patterns in vast data sets, deep learning models have revolutionized predictive modeling, often achieving superior performance compared to standard mathematical and statistical models for infectious disease forecasting. Our proposed models yielded highly accurate results in this setting, with incorporation of social media data offering mild to moderate increases in forecasting accuracy. Moreover, the incorporation of social media data helps capture trends in geographic regions that may be underrepresented in traditional health records. However, continuous refinement of modeling frameworks such as the one presented here is necessary due to the constantly changing nature of data sources.

## Data Availability

All data produced in the present study are available upon reasonable request to the authors

https://github.com/nytimes/covid-19-data/tree/master

## Ethics statement

Ethical review for this study was obtained by the Institutional Review Board of Clemson University (#2020-0150). No consent was needed for this study; retrospective data based on medical claims, electronic health records, and digital trace data were de-identified to study investigators.

## Author Statement

LR and JW conceptualized this study. JW conducted all data analyses and prepared the original draft of the manuscript, with contributions from LR. All authors contributed to the revised manuscript. ST and JW led data visualization. JW developed the methodology for this study, with contributions from MW. JW and TA accessed and verified the data underlying this study. All authors interpreted study findings. LR performed project administration, acquired funding, and supervised this study. All authors contributed to reviewing and editing the work.

## Acknowledgements

This project has been funded by the National Library of Medicine of the National Institutes of Health (NIH) under award number R01LM014193 and the Center for Forecasting and Outbreak Analytics of the Centers for Disease Control and Prevention (CDC) under award number NU38FT000011. We thank Dr. Fatih Gezer and Dr. Sakhawat Hossain for their assistance with data-related questions. We thank Clemson University’s Social Media Listening Center for facilitating the procurement of social media data. The content and decision to publish is solely based on the authors of this study and does not necessarily represent the official views of the NIH or CDC. The funders had no role in the study design, data collection and analysis, decision to publish, or preparation of this manuscript.

