## Supplementary material for "Enhancing Pandemic Prediction: A Deep Learning Approach Using Transformer Neural Networks and Multi-Source Data Fusion for Infectious Disease Forecasting": Table A

**Table A1. County-Level Root Mean Square Error (RMSE) and interquartile range (IQR) for daily prediction (Full model).**

| | | RMSE: day 1 prediction | RMSE: day 2 prediction | RMSE: day 3 prediction | RMSE: day 4 prediction | RMSE: day 5 prediction | RMSE: day 6 prediction | RMSE: day 7 prediction |
|---|---|---|---|---|---|---|---|---|
| **cases** | | | | | | | | |
| 12/01/2021 - | Median | 7.6 | 10.01 | 10.77 | 11.14 | 13.26 | 9.8 | 10.22 |
| 3/31/22 | (Q1; Q3) | (3.27; 17.24) | (4.64; 22.15) | (5.06; 24.27) | (5.21; 28.14) | (6.11; 33.77) | (4.43; 21.98) | (4.74; 21.97) |
| 05/01/2022 - | Median | 2.91 | 3.24 | 3.1 | 3.4 | 4.1 | 2.87 | 2.91 |
| 10/31/22 | (Q1; Q3) | (0.94; 4.19) | (0.97; 4.65) | (0.94; 4.44) | (1; 4.85) | (1.2; 6.64) | (0.98; 4.83) | (0.99; 4.8) |
| 12/01/2022 - | Median | 1.61 | 1.59 | 1.54 | 1.82 | 2.34 | 1.61 | 1.58 |
| 2/28/23 | (Q1; Q3) | (0.87; 4.28) | (0.86; 4.37) | (0.83; 4.18) | (0.97; 5) | (1.23; 6.79) | (0.84; 4.24) | (0.84; 4.2) |
| **deaths** | | | | | | | | |
| 12/01/2021 - | Median | 0.05 | 0.05 | 0.05 | 0.05 | 0.06 | 0.05 | 0.05 |
| 3/31/22 | (Q1; Q3) | (0.03; 0.1) | (0.02; 0.09) | (0.02; 0.09) | (0.02; 0.09) | (0.03; 0.11) | (0.02; 0.09) | (0.03; 0.1) |
| 05/01/2022 - | Median | 0.02 | 0.02 | 0.02 | 0.02 | 0.02 | 0.02 | 0.02 |
| 10/31/22 | (Q1; Q3) | (0.01; 0.03) | (0.01; 0.02) | (0.01; 0.03) | (0.01; 0.02) | (0.01; 0.03) | (0.01; 0.03) | (0.01; 0.03) |
| 12/01/2022 - | Median | 0.01 | 0.01 | 0.01 | 0.01 | 0.01 | 0.01 | 0.01 |
| 2/28/23 | (Q1; Q3) | (0.01; 0.03) | (0.01; 0.02) | (0.01; 0.03) | (0.01; 0.02) | (0.01; 0.02) | (0.01; 0.03) | (0.01; 0.03) |

**Note: RMSE: day x prediction: County level RMSE for x days ahead prediction; Q1: First Quartile; Q3: Third Quartile.**

**Table A2. County-Level Root Mean Square Error (RMSE) and interquartile range (IQR) for weekly prediction (Full model).**

| | | RMSE: week 1 prediction | RMSE: week 2 prediction | RMSE: week 3 prediction | RMSE: week 4 prediction | RMSE: week 5 prediction |
|---|---|---|---|---|---|---|
| **cases** | | | | | | |
| 12/01/2021 - | Median | 146.55 | 145.66 | 151.35 | 157.76 | 167.74 |
| 3/31/22 | (Q1; Q3) | (67.34; 356.04) | (68.78; 374.51) | (73.59; 390.75) | (77.56; 426.13) | (83.83; 462.56) |
| 05/01/2022 - | Median | 31.89 | 32.31 | 34.55 | 37.94 | 42.47 |
| 10/31/22 | (Q1; Q3) | (10.9; 51.26) | (11.06; 50.73) | (11.5; 51.91) | (11.86; 54.89) | (12.84; 61.6) |
| 12/01/2022 - | Median | 18.7 | 18.27 | 19.06 | 20.86 | 23.3 |
| 2/28/23 | (Q1; Q3) | (9.63; 46.65) | (9.68; 44.81) | (10.37; 46.02) | (11.13; 50.6) | (12.29; 59.46) |
| **deaths** | | | | | | |
| 12/01/2021 - | Median | 0.59 | 0.56 | 0.52 | 0.54 | 0.54 |
| 3/31/22 | (Q1; Q3) | (0.29; 1.18) | (0.27; 1.05) | (0.26; 0.94) | (0.25; 0.94) | (0.24; 0.96) |
| 05/01/2022 - | Median | 0.17 | 0.18 | 0.17 | 0.16 | 0.16 |
| 10/31/22 | (Q1; Q3) | (0.1; 0.35) | (0.1; 0.36) | (0.1; 0.29) | (0.09; 0.3) | (0.09; 0.32) |
| 12/01/2022 - | Median | 0.11 | 0.13 | 0.11 | 0.11 | 0.12 |
| 2/28/23 | (Q1; Q3) | (0.06; 0.26) | (0.06; 0.29) | (0.06; 0.24) | (0.06; 0.26) | (0.06; 0.27) |

**Note: RMSE: week x prediction: County level RMSE for x weeks ahead prediction; Q1: First Quartile; Q3: Third Quartile.**

**Table A3. County-Level Mean Absolute Error (MAE) and interquartile range (IQR) for daily prediction (Full model).**

| | | MAE: day 1 prediction | MAE: day 2 prediction | MAE: day 3 prediction | MAE: day 4 prediction | MAE: day 5 prediction | MAE: day 6 prediction | MAE: day 7 prediction |
|---|---|---|---|---|---|---|---|---|
| **cases** | | | | | | | | |
| 12/01/2021 - | Median | 4.04 | 4.85 | 5.3 | 5.93 | 7.5 | 5.53 | 5.72 |
| 3/31/22 | (Q1; Q3) | (1.77; 9.29) | (2.3; 10.2) | (2.46; 12.43) | (2.71; 16.03) | (3.44; 20.15) | (2.49; 12.33) | (2.66; 12.4) |
| 05/01/2022 - | Median | 1.41 | 1.12 | 1.07 | 1.57 | 2.22 | 1.52 | 1.48 |
| 10/31/22 | (Q1; Q3) | (0.53; 2.51) | (0.41; 2.02) | (0.37; 1.6) | (0.54; 3.03) | (0.8; 4.59) | (0.63; 3.4) | (0.62; 3.33) |
| 12/01/2022 - | Median | 1.06 | 0.71 | 0.58 | 1.19 | 1.84 | 1.06 | 0.98 |
| 2/28/23 | (Q1; Q3) | (0.53; 3.19) | (0.4; 2.18) | (0.32; 1.63) | (0.58; 3.37) | (0.89; 5.66) | (0.54; 2.96) | (0.5; 2.66) |
| **deaths** | | | | | | | | |
| 12/01/2021 - | Median | 0.03 | 0.02 | 0.03 | 0.03 | 0.04 | 0.04 | 0.04 |
| 3/31/22 | (Q1; Q3) | (0.01; 0.06) | (0.01; 0.05) | (0.02; 0.06) | (0.01; 0.05) | (0.01; 0.08) | (0.02; 0.07) | (0.02; 0.08) |
| 05/01/2022 - | Median | 0.01 | 0.01 | 0.01 | 0.01 | 0.01 | 0.01 | 0.01 |
| 10/31/22 | (Q1; Q3) | (0; 0.01) | (0; 0.01) | (0; 0.02) | (0; 0.01) | (0; 0.01) | (0; 0.01) | (0; 0.01) |
| 12/01/2022 - | Median | 0.01 | 0.01 | 0.01 | 0.01 | 0.01 | 0.01 | 0.01 |
| 2/28/23 | (Q1; Q3) | (0; 0.01) | (0; 0.01) | (0; 0.02) | (0; 0.01) | (0; 0.01) | (0; 0.02) | (0; 0.02) |

**Note: MAE: day x prediction: County level MAE for x days ahead prediction; Q1: First Quartile; Q3: Third Quartile.**

**Table A4. County-Level Mean Absolute Error (MAE) and interquartile range (IQR) for weekly prediction (Full model).**

| | | MAE: week 1 prediction | MAE: week 2 prediction | MAE: week 3 prediction | MAE: week 4 prediction | MAE: week 5 prediction |
|---|---|---|---|---|---|---|
| **cases** | | | | | | |
| 12/01/2021 - | Median | 91.57 | 93.14 | 96.86 | 100.7 | 111.94 |
| 3/31/22 | (Q1; Q3) | (43.44; 217.28) | (44.36; 237.99) | (47.17; 257.68) | (50.15; 288.72) | (54.93; 318.45) |
| 05/01/2022 - | Median | 21.54 | 21.81 | 22.52 | 23.51 | 26.32 |
| 10/31/22 | (Q1; Q3) | (8.32; 38) | (8.33; 37.38) | (8.67; 38.52) | (8.82; 40.61) | (9.7; 45.74) |
| 12/01/2022 - | Median | 15.29 | 14.66 | 14.43 | 15.7 | 18.02 |
| 2/28/23 | (Q1; Q3) | (7.42; 36.43) | (7.21; 35.16) | (7.56; 35.71) | (8.48; 38.48) | (10.05; 47.94) |
| **deaths** | | | | | | |
| 12/01/2021 - | Median | 0.41 | 0.38 | 0.37 | 0.38 | 0.41 |
| 3/31/22 | (Q1; Q3) | (0.18; 0.8) | (0.16; 0.72) | (0.15; 0.65) | (0.16; 0.64) | (0.16; 0.67) |
| 05/01/2022 - | Median | 0.1 | 0.1 | 0.08 | 0.08 | 0.09 |
| 10/31/22 | (Q1; Q3) | (0.04; 0.19) | (0.04; 0.2) | (0.04; 0.18) | (0.04; 0.18) | (0.04; 0.2) |
| 12/01/2022 - | Median | 0.06 | 0.08 | 0.08 | 0.06 | 0.06 |
| 2/28/23 | (Q1; Q3) | (0.03; 0.15) | (0.03; 0.19) | (0.03; 0.17) | (0.03; 0.15) | (0.03; 0.16) |

**Note: MAE: week x prediction: County level MAE for x weeks ahead prediction; Q1: First Quartile; Q3: Third Quartile.**

**Table A5. County-Level Percentage Agreement (PA %) and interquartile range (IQR) for daily prediction (Full model trained with no X input).**

| | | PA: day 1 prediction | PA: day 2 prediction | PA: day 3 prediction | PA: day 4 prediction | PA: day 5 prediction | PA: day 6 prediction | PA: day 7 prediction |
|---|---|---|---|---|---|---|---|---|
| **cases** | | | | | | | | |
| 12/01/2021 - | Median | 79.56 | 76.16 | 75.89 | 75.84 | 75.65 | 79.89 | 77.89 |
| 3/31/22 | (Q1; Q3) | (77.79; 81.09) | (74.75; 78.05) | (74.18; 77.61) | (73.08; 77.88) | (73.38; 77.75) | (77.95; 82.01) | (76.3; 79.88) |
| 05/01/2022 - | Median | 81.06 | 74.65 | 74.78 | 72.83 | 74.29 | 74.58 | 72.13 |
| 10/31/22 | (Q1; Q3) | (76.98; 82.91) | (69.54; 77.25) | (69.89; 77.66) | (68.34; 76.46) | (69.26; 76.94) | (69.77; 78.32) | (67.02; 75.09) |
| 12/01/2022 - | Median | 81.5 | 75.16 | 76.24 | 75.89 | 75.75 | 77.06 | 74.83 |
| 2/28/23 | (Q1; Q3) | (77.39; 83.44) | (72.33; 77.5) | (72.5; 78.18) | (72.13; 78.93) | (72.53; 78.68) | (74.47; 79.72) | (72.19; 76.79) |
| **deaths** | | | | | | | | |
| 12/01/2021 - | Median | 82.1 | 83.2 | 82.84 | 82.26 | 81.88 | 79.03 | 78.66 |
| 3/31/22 | (Q1; Q3) | (69.91; 85.86) | (70.28; 86.29) | (71.56; 86.35) | (70.65; 85.8) | (71.59; 85.72) | (65.47; 84.56) | (65.07; 83.48) |
| 05/01/2022 - | Median | 57.64 | 58.22 | 58.84 | 59.75 | 59.2 | 56.93 | 56.73 |
| 10/31/22 | (Q1; Q3) | (37.16; 69.8) | (40; 70.19) | (35.83; 69.87) | (40; 69.83) | (40; 70.78) | (38.75; 68.26) | (38.75; 65.63) |
| 12/01/2022 - | Median | 67.62 | 67.72 | 67.88 | 67.9 | 67.85 | 62.7 | 61.6 |
| 2/28/23 | (Q1; Q3) | (41.67; 76.18) | (39.65; 79.05) | (41.67; 77.82) | (41.67; 76.93) | (41.67; 76.85) | (41.67; 73.09) | (41.67; 71.38) |

**Note: PA: day x prediction: County level PA for x days ahead prediction; Q1: First Quartile; Q3: Third Quartile.**

**Table A6. County-Level Percentage Agreement (PA %) and interquartile range (IQR) for weekly prediction (Full model trained with no X input).**

| | | PA: week 1 prediction | PA: week 2 prediction | PA: week 3 prediction | PA: week 4 prediction | PA: week 5 prediction |
|---|---|---|---|---|---|---|
| **cases** | | | | | | |
| 12/01/2021 - | Median | 64.43 | 63.88 | 64.09 | 64.06 | 63.02 |
| 3/31/22 | (Q1; Q3) | (62.79; 66.25) | (62.34; 65.92) | (62.2; 65.98) | (61.54; 65.81) | (61.19; 65.55) |
| 05/01/2022 - | Median | 80.43 | 78.16 | 76.94 | 73.53 | 72.94 |
| 10/31/22 | (Q1; Q3) | (76.65; 83.29) | (73.89; 81.07) | (72.36; 79.62) | (68.99; 76.16) | (68.66; 75.27) |
| 12/01/2022 - | Median | 78.69 | 77.13 | 76 | 74.72 | 73.35 |
| 2/28/23 | (Q1; Q3) | (75.09; 81.23) | (73.21; 79.64) | (73.24; 79.46) | (71.12; 78.4) | (69.99; 77.08) |
| **deaths** | | | | | | |
| 12/01/2021 - | Median | 83.58 | 83.6 | 83.08 | 82.22 | 82.75 |
| 3/31/22 | (Q1; Q3) | (81.13; 86.07) | (80.39; 86.66) | (79.77; 85.48) | (78.81; 84.86) | (79.59; 85.04) |
| 05/01/2022 - | Median | 80.39 | 83.74 | 82.42 | 80.15 | 82.63 |
| 10/31/22 | (Q1; Q3) | (76.75; 82.47) | (81.15; 85.62) | (80.09; 85.17) | (77.8; 83.17) | (79.51; 84.43) |
| 12/01/2022 - | Median | 81.79 | 85.93 | 84.37 | 81.76 | 83.5 |
| 2/28/23 | (Q1; Q3) | (77.81; 85.12) | (81.67; 88.42) | (79.23; 87.22) | (77.11; 85.04) | (79.09; 87.26) |

**Note: PA: week x prediction: County level PA for x weeks ahead prediction; Q1: First Quartile; Q3: Third Quartile.**

Table A7. County-Level Root Mean Square Error (RMSE) and interquartile range (IQR) for daily prediction (Full model trained with no X input).

| | | RMSE: day 1 prediction | RMSE: day 2 prediction | RMSE: day 3 prediction | RMSE: day 4 prediction | RMSE: day 5 prediction | RMSE: day 6 prediction | RMSE: day 7 prediction |
|---|---|---|---|---|---|---|---|---|
| **cases** | | | | | | | | |
| 12/01/2021 - | Median | 14.43 | 15.54 | 16.14 | 15.18 | 15.49 | 13.69 | 16.38 |
| 3/31/22 | (Q1; Q3) | (6.38; 35.04) | (7.2; 37.53) | (7.7; 37.98) | (7.64; 36.7) | (7.48; 37.22) | (6.58; 33.56) | (7.8; 40.11) |
| 05/01/2022 - | Median | 3.18 | 4.13 | 4.6 | 4.66 | 4.96 | 3.84 | 4.61 |
| 10/31/22 | (Q1; Q3) | (1.08; 6.4) | (1.45; 7.92) | (1.57; 6.85) | (1.55; 6.76) | (1.63; 7.14) | (1.32; 6.49) | (1.58; 8.43) |
| 12/01/2022 - | Median | 1.77 | 2.27 | 2.48 | 2.5 | 2.68 | 2.2 | 2.62 |
| 2/28/23 | (Q1; Q3) | (0.96; 4.42) | (1.21; 5.96) | (1.31; 6.36) | (1.34; 6.27) | (1.53; 6.87) | (1.15; 5.76) | (1.34; 7) |
| **deaths** | | | | | | | | |
| 12/01/2021 - | Median | 0.08 | 0.07 | 0.07 | 0.07 | 0.07 | 0.07 | 0.08 |
| 3/31/22 | (Q1; Q3) | (0.04; 0.14) | (0.03; 0.12) | (0.03; 0.12) | (0.03; 0.11) | (0.03; 0.12) | (0.03; 0.13) | (0.03; 0.14) |
| 05/01/2022 - | Median | 0.03 | 0.03 | 0.02 | 0.02 | 0.02 | 0.02 | 0.02 |
| 10/31/22 | (Q1; Q3) | (0.02; 0.04) | (0.01; 0.03) | (0.01; 0.03) | (0.01; 0.03) | (0.01; 0.03) | (0.01; 0.04) | (0.01; 0.04) |
| 12/01/2022 - | Median | 0.02 | 0.02 | 0.01 | 0.01 | 0.01 | 0.02 | 0.02 |
| 2/28/23 | (Q1; Q3) | (0.01; 0.04) | (0.01; 0.04) | (0.01; 0.03) | (0.01; 0.04) | (0.01; 0.04) | (0.01; 0.04) | (0.01; 0.04) |

**Note: RMSE: day x prediction: County level RMSE for x days ahead prediction; Q1: First Quartile; Q3: Third Quartile.**

**Table A8. County-Level Root Mean Square Error (RMSE) and interquartile range (IQR) for weekly prediction (Full model trained with no X input).**

| | | RMSE: week 1 prediction | RMSE: week 2 prediction | RMSE: week 3 prediction | RMSE: week 4 prediction | RMSE: week 5 prediction |
|---|---|---|---|---|---|---|
| **cases** | | | | | | |
| 12/01/2021 - | Median | 215.7 | 214.67 | 216.11 | 212.64 | 213.63 |
| 3/31/22 | (Q1; Q3) | (103.14; 559.54) | (104.43; 562.79) | (104.9; 556.36) | (103.3; 549.05) | (102.74; 556.65) |
| 05/01/2022 - | Median | 36.14 | 40.67 | 44.17 | 48.13 | 52.79 |
| 10/31/22 | (Q1; Q3) | (11.3; 60.52) | (13.62; 65.52) | (14.8; 68.82) | (16.32; 73.89) | (18.12; 79.6) |
| 12/01/2022 - | Median | 20.69 | 23.72 | 25.37 | 27.33 | 29.64 |
| 2/28/23 | (Q1; Q3) | (11.7; 50.53) | (12.91; 57.25) | (13.62; 60.92) | (14.56; 65.11) | (16.38; 71.38) |
| **deaths** | | | | | | |
| 12/01/2021 - | Median | 0.79 | 0.8 | 0.75 | 0.74 | 0.72 |
| 3/31/22 | (Q1; Q3) | (0.34; 1.3) | (0.36; 1.28) | (0.36; 1.27) | (0.36; 1.3) | (0.36; 1.27) |
| 05/01/2022 - | Median | 0.26 | 0.23 | 0.23 | 0.22 | 0.22 |
| 10/31/22 | (Q1; Q3) | (0.14; 0.43) | (0.13; 0.41) | (0.12; 0.42) | (0.12; 0.44) | (0.13; 0.44) |
| 12/01/2022 - | Median | 0.16 | 0.12 | 0.12 | 0.14 | 0.13 |
| 2/28/23 | (Q1; Q3) | (0.08; 0.4) | (0.07; 0.36) | (0.07; 0.37) | (0.07; 0.39) | (0.07; 0.38) |

**Note: RMSE: week x prediction: County level RMSE for x weeks ahead prediction; Q1: First Quartile; Q3: Third Quartile.**

**Table A9. County-Level Mean Absolute Error (MAE) and interquartile range (IQR) for daily prediction (Full model trained with no X input).**

| | | MAE: day 1 prediction | MAE: day 2 prediction | MAE: day 3 prediction | MAE: day 4 prediction | MAE: day 5 prediction | MAE: day 6 prediction | MAE: day 7 prediction |
|---|---|---|---|---|---|---|---|---|
| **cases** | | | | | | | | |
| 12/01/2021 - | Median | 8.2 | 8.62 | 8.66 | 8.44 | 8.96 | 8 | 9.62 |
| 3/31/22 | (Q1; Q3) | (3.59; 20.49) | (4.11; 21.95) | (4.2; 21.05) | (4.05; 20.82) | (4.3; 22.13) | (3.64; 18.31) | (4.54; 23.1) |
| 05/01/2022 - | Median | 1.97 | 2.44 | 2.2 | 2.2 | 2.58 | 2.19 | 2.84 |
| 10/31/22 | (Q1; Q3) | (0.77; 4.11) | (0.95; 5.03) | (0.82; 4.1) | (0.82; 4.01) | (0.97; 4.31) | (0.89; 4.85) | (1.19; 6.75) |
| 12/01/2022 - | Median | 1.17 | 1.49 | 1.44 | 1.31 | 1.67 | 1.58 | 2.05 |
| 2/28/23 | (Q1; Q3) | (0.6; 3.35) | (0.76; 4.29) | (0.73; 3.78) | (0.72; 3.51) | (0.92; 4.4) | (0.75; 4.27) | (1; 5.72) |
| **deaths** | | | | | | | | |
| 12/01/2021 - | Median | 0.05 | 0.04 | 0.04 | 0.04 | 0.04 | 0.05 | 0.06 |
| 3/31/22 | (Q1; Q3) | (0.02; 0.1) | (0.02; 0.08) | (0.01; 0.07) | (0.01; 0.07) | (0.02; 0.08) | (0.02; 0.11) | (0.03; 0.12) |
| 05/01/2022 - | Median | 0.01 | 0.01 | 0 | 0.01 | 0.01 | 0.01 | 0.01 |
| 10/31/22 | (Q1; Q3) | (0.01; 0.03) | (0; 0.02) | (0; 0.01) | (0; 0.01) | (0; 0.02) | (0.01; 0.02) | (0.01; 0.03) |
| 12/01/2022 - | Median | 0.01 | 0.01 | 0 | 0.01 | 0.01 | 0.01 | 0.01 |
| 2/28/23 | (Q1; Q3) | (0; 0.04) | (0; 0.02) | (0; 0.02) | (0; 0.02) | (0; 0.02) | (0.01; 0.03) | (0.01; 0.03) |

**Note: MAE: day x prediction: County level MAE for x days ahead prediction; Q1: First Quartile; Q3: Third Quartile.**

**Table A10. County-Level Mean Absolute Error (MAE) and interquartile range (IQR) for weekly prediction (Full model trained with no X input).**

| | | MAE: week 1 prediction | MAE: week 2 prediction | MAE: week 3 prediction | MAE: week 4 prediction | MAE: week 5 prediction |
|---|---|---|---|---|---|---|
| **cases** | | | | | | |
| 12/01/2021 - | Median | 139.09 | 140.17 | 140.33 | 137.09 | 136.32 |
| 3/31/22 | (Q1; Q3) | (67.16; 365.43) | (69.39; 367.08) | (68.94; 359.12) | (67.62; 350.21) | (68.42; 364.77) |
| 05/01/2022 - | Median | 23.49 | 28 | 30.67 | 34.19 | 37.36 |
| 10/31/22 | (Q1; Q3) | (8.31; 42.9) | (10.26; 48.16) | (11.41; 49.84) | (12.33; 57.05) | (13.56; 60.19) |
| 12/01/2022 - | Median | 16.7 | 18.62 | 19.46 | 20.81 | 22.23 |
| 2/28/23 | (Q1; Q3) | (8.7; 38.95) | (9.82; 43.56) | (10.21; 46.73) | (10.91; 50.05) | (12.05; 55.14) |
| **deaths** | | | | | | |
| 12/01/2021 - | Median | 0.52 | 0.53 | 0.5 | 0.51 | 0.53 |
| 3/31/22 | (Q1; Q3) | (0.2; 0.94) | (0.19; 0.9) | (0.2; 0.92) | (0.21; 0.96) | (0.21; 0.94) |
| 05/01/2022 - | Median | 0.11 | 0.1 | 0.1 | 0.11 | 0.11 |
| 10/31/22 | (Q1; Q3) | (0.06; 0.27) | (0.05; 0.22) | (0.05; 0.23) | (0.05; 0.25) | (0.05; 0.25) |
| 12/01/2022 - | Median | 0.11 | 0.07 | 0.07 | 0.1 | 0.08 |
| 2/28/23 | (Q1; Q3) | (0.04; 0.25) | (0.03; 0.21) | (0.03; 0.23) | (0.04; 0.26) | (0.03; 0.24) |

**Note: MAE: week x prediction: County level MAE for x weeks ahead prediction; Q1: First Quartile; Q3: Third Quartile.**

Table A11. County-Level Percentage Agreement (PA %) and interquartile range (IQR) for daily prediction (Persistence model).

| | | PA: day 1 prediction | PA: day 2 prediction | PA: day 3 prediction | PA: day 4 prediction | PA: day 5 prediction | PA: day 6 prediction | PA: day 7 prediction |
|---|---|---|---|---|---|---|---|---|
| **cases** | | | | | | | | |
| 12/01/2021 - | Median | 85.47 | 78.78 | 73.46 | 70.33 | 67.47 | 64.3 | 60.25 |
| 3/31/22 | (Q1; Q3) | (83.11; 87.12) | (76.5; 80.2) | (71; 75.47) | (67.8; 71.97) | (64.72; 68.85) | (61.42; 65.91) | (57.45; 62.04) |
| 05/01/2022 - | Median | 76.04 | 76.04 | 76.04 | 74.89 | 74.89 | 75.54 | 73.02 |
| 10/31/22 | (Q1; Q3) | (71.75; 79.29) | (71.75; 79.29) | (71.75; 79.29) | (69.93; 78.09) | (69.93; 78.09) | (68.86; 77.85) | (67.32; 76.64) |
| 12/01/2022 - | Median | 77.97 | 77.97 | 76.23 | 76.23 | 76.23 | 76.23 | 72.58 |
| 2/28/23 | (Q1; Q3) | (74.87; 80.6) | (74.87; 80.6) | (73.22; 78.25) | (73.22; 78.25) | (73.22; 78.25) | (73.22; 78.25) | (68.97; 75.12) |
| **deaths** | | | | | | | | |
| 12/01/2021 - | Median | 84.98 | 82.84 | 80.99 | 80.03 | 77.75 | 76.9 | 74.67 |
| 3/31/22 | (Q1; Q3) | (75.01; 88.83) | (72.17; 87) | (69.31; 85.5) | (67.36; 84.71) | (67.48; 83.85) | (66.18; 82.59) | (65.18; 81.23) |
| 05/01/2022 - | Median | 59.42 | 59.42 | 60.04 | 58 | 58 | 59.42 | 59.42 |
| 10/31/22 | (Q1; Q3) | (40; 70.65) | (40; 70.65) | (40; 70.65) | (43.33; 68.54) | (43.33; 68.54) | (40.25; 68.09) | (40.25; 67.56) |
| 12/01/2022 - | Median | 68.82 | 68.82 | 68.06 | 68.06 | 68.06 | 68.06 | 67.32 |
| 2/28/23 | (Q1; Q3) | (41.67; 79.25) | (41.67; 79.25) | (41.67; 78.8) | (41.67; 78.8) | (44.44; 78.8) | (44.44; 78.8) | (44.44; 77.87) |

Note: PA: day x prediction: County level PA for x days ahead prediction; Q1: First Quartile; Q3: Third Quartile.

Table A12. County-Level Percentage Agreement (PA %) and interquartile range (IQR) for weekly prediction (Persistence model).

| | | PA: week 1 prediction | PA: week 2 prediction | PA: week 3 prediction | PA: week 4 prediction | PA: week 5 prediction |
|---|---|---|---|---|---|---|
| **cases** | | | | | | |
| 12/01/2021 - | Median | 64.16 | 47.48 | 36.24 | 30.52 | 26.85 |
| 3/31/22 | (Q1; Q3) | (62.48; 66.54) | (44.77; 49.44) | (34.06; 39.51) | (28.55; 33.01) | (24.95; 29.08) |
| 05/01/2022 - | Median | 77.08 | 67.91 | 59.34 | 54.23 | 49.45 |
| 10/31/22 | (Q1; Q3) | (72.4; 79.76) | (63.15; 70.08) | (56.99; 62.96) | (50.9; 57.14) | (44.94; 52.05) |
| 12/01/2022 - | Median | 76.08 | 70.62 | 62.7 | 58.58 | 55.2 |
| 2/28/23 | (Q1; Q3) | (73.45; 79.39) | (65.96; 73.62) | (58.94; 66.2) | (54.34; 62.86) | (50.34; 59) |
| **deaths** | | | | | | |
| 12/01/2021 - | Median | 83.82 | 73.67 | 65.9 | 58.97 | 53 |
| 3/31/22 | (Q1; Q3) | (81.17; 86.18) | (68.7; 76.88) | (59.31; 69.4) | (51.58; 63.42) | (46.72; 59.54) |
| 05/01/2022 - | Median | 83.3 | 72.86 | 63.31 | 54.94 | 48.72 |
| 10/31/22 | (Q1; Q3) | (82.02; 86.2) | (68.72; 76.31) | (57.41; 69.87) | (50.92; 65.25) | (43.27; 60.43) |
| 12/01/2022 - | Median | 84.42 | 74.02 | 65.09 | 60.05 | 57.15 |
| 2/28/23 | (Q1; Q3) | (78.57; 89.09) | (65.31; 81.85) | (57.14; 75.94) | (47.98; 71.33) | (44.46; 69.72) |

Note: PA: week x prediction: County level PA for x weeks ahead prediction; Q1: First Quartile; Q3: Third Quartile.

**Table A13. County-Level Root Mean Square Error (RMSE) and interquartile range (IQR) for daily prediction (Persistence model).**

| | | RMSE: day 1 prediction | RMSE: day 2 prediction | RMSE: day 3 prediction | RMSE: day 4 prediction | RMSE: day 5 prediction | RMSE: day 6 prediction | RMSE: day 7 prediction |
|---|---|---|---|---|---|---|---|---|
| **cases** | | | | | | | | |
| 12/01/2021 - | Median | 8.39 | 13.19 | 17 | 21.03 | 24.65 | 28.36 | 32.37 |
| 3/31/22 | (Q1; Q3) | (3.86; 19.5) | (6.27; 31.93) | (8.33; 42.07) | (10.13; 51.99) | (11.87; 61.74) | (13.76; 71.79) | (15.74; 82.2) |
| 05/01/2022 - | Median | 2.68 | 3.8 | 4.64 | 5.36 | 6 | 6.53 | 6.86 |
| 10/31/22 | (Q1; Q3) | (0.88; 4.05) | (1.23; 5.72) | (1.52; 7) | (1.75; 8.09) | (1.96; 9.06) | (2.12; 9.73) | (2.27; 10.38) |
| 12/01/2022 - | Median | 1.46 | 2.05 | 2.51 | 2.91 | 3.25 | 3.56 | 3.84 |
| 2/28/23 | (Q1; Q3) | (0.78; 3.8) | (1.1; 5.15) | (1.36; 6.26) | (1.57; 7.2) | (1.76; 8.03) | (1.92; 8.78) | (2.06; 9.3) |
| **deaths** | | | | | | | | |
| 12/01/2021 - | Median | 0.04 | 0.06 | 0.08 | 0.09 | 0.1 | 0.11 | 0.12 |
| 3/31/22 | (Q1; Q3) | (0.02; 0.07) | (0.03; 0.11) | (0.03; 0.13) | (0.04; 0.16) | (0.04; 0.18) | (0.05; 0.19) | (0.05; 0.21) |
| 05/01/2022 - | Median | 0.01 | 0.02 | 0.03 | 0.03 | 0.03 | 0.04 | 0.04 |
| 10/31/22 | (Q1; Q3) | (0.01; 0.02) | (0.01; 0.03) | (0.01; 0.04) | (0.02; 0.05) | (0.02; 0.06) | (0.02; 0.06) | (0.02; 0.07) |
| 12/01/2022 - | Median | 0.01 | 0.01 | 0.02 | 0.02 | 0.02 | 0.02 | 0.02 |
| 2/28/23 | (Q1; Q3) | (0.01; 0.02) | (0.01; 0.03) | (0.01; 0.04) | (0.01; 0.04) | (0.01; 0.05) | (0.01; 0.05) | (0.01; 0.05) |

**Note: RMSE: day x prediction: County level RMSE for x days ahead prediction; Q1: First Quartile; Q3: Third Quartile.**

**Table A14. County-Level Root Mean Square Error (RMSE) and interquartile range (IQR) for weekly prediction (Persistence model).**

| | | RMSE: week 1 prediction | RMSE: week 2 prediction | RMSE: week 3 prediction | RMSE: week 4 prediction | RMSE: week 5 prediction |
|---|---|---|---|---|---|---|
| **cases** | | | | | | |
| 12/01/2021 - | Median | 213.88 | 389.37 | 528.54 | 615.66 | 665.7 |
| 3/31/22 | (Q1; Q3) | (103.9; 556.19) | (180.68; 1005.83) | (241.81; 1342.58) | (284.74; 1577.62) | (309.56; 1718.86) |
| 05/01/2022 - | Median | 43.89 | 58.34 | 66.79 | 74.35 | 82.82 |
| 10/31/22 | (Q1; Q3) | (14.71; 68.35) | (20.22; 92.57) | (24.52; 110.12) | (27.82; 138.24) | (29.61; 162.66) |
| 12/01/2022 - | Median | 25.33 | 32.99 | 41.22 | 48.22 | 53.28 |
| 2/28/23 | (Q1; Q3) | (13.63; 60.52) | (17.84; 77.79) | (22.06; 94.36) | (23.45; 117.36) | (24.84; 127.18) |
| **deaths** | | | | | | |
| 12/01/2021 - | Median | 0.74 | 1.06 | 1.44 | 1.69 | 1.94 |
| 3/31/22 | (Q1; Q3) | (0.35; 1.27) | (0.55; 2.13) | (0.73; 2.88) | (0.86; 3.51) | (0.93; 4.2) |
| 05/01/2022 - | Median | 0.23 | 0.38 | 0.5 | 0.6 | 0.72 |
| 10/31/22 | (Q1; Q3) | (0.14; 0.41) | (0.23; 0.88) | (0.3; 1.11) | (0.36; 1.28) | (0.45; 1.51) |
| 12/01/2022 - | Median | 0.13 | 0.2 | 0.23 | 0.28 | 0.32 |
| 2/28/23 | (Q1; Q3) | (0.07; 0.36) | (0.11; 0.59) | (0.13; 0.72) | (0.16; 0.8) | (0.21; 0.88) |

**Note: RMSE: week x prediction: County level RMSE for x weeks ahead prediction; Q1: First Quartile; Q3: Third Quartile.**

**Table A15. County-Level Mean Absolute Error (MAE) and interquartile range (IQR) for daily prediction (Persistence model).**

| | | MAE: day 1 prediction | MAE: day 2 prediction | MAE: day 3 prediction | MAE: day 4 prediction | MAE: day 5 prediction | MAE: day 6 prediction | MAE: day 7 prediction |
|---|---|---|---|---|---|---|---|---|
| **cases** | | | | | | | | |
| 12/01/2021 - | Median | 3.62 | 6.74 | 9.69 | 12.51 | 15.22 | 17.86 | 20.58 |
| 3/31/22 | (Q1; Q3) | (1.67; 8.21) | (3.09; 15.66) | (4.46; 23.28) | (5.75; 30.77) | (7.11; 38.14) | (8.39; 45.67) | (9.7; 52.99) |
| 05/01/2022 - | Median | 0.66 | 1.32 | 1.98 | 2.64 | 3.3 | 3.92 | 4.48 |
| 10/31/22 | (Q1; Q3) | (0.25; 1.08) | (0.5; 2.15) | (0.75; 3.23) | (1; 4.34) | (1.25; 5.44) | (1.47; 6.44) | (1.67; 7.41) |
| 12/01/2022 - | Median | 0.41 | 0.81 | 1.23 | 1.64 | 2.05 | 2.46 | 2.83 |
| 2/28/23 | (Q1; Q3) | (0.22; 1.15) | (0.44; 2.16) | (0.66; 3.18) | (0.87; 4.22) | (1.09; 5.26) | (1.31; 6.31) | (1.52; 7.15) |
| **deaths** | | | | | | | | |
| 12/01/2021 - | Median | 0.01 | 0.03 | 0.04 | 0.05 | 0.06 | 0.07 | 0.08 |
| 3/31/22 | (Q1; Q3) | (0; 0.03) | (0.01; 0.05) | (0.01; 0.07) | (0.02; 0.09) | (0.02; 0.11) | (0.02; 0.13) | (0.03; 0.14) |
| 05/01/2022 - | Median | 0 | 0 | 0.01 | 0.01 | 0.01 | 0.01 | 0.01 |
| 10/31/22 | (Q1; Q3) | (0; 0) | (0; 0.01) | (0; 0.01) | (0; 0.02) | (0.01; 0.02) | (0.01; 0.03) | (0.01; 0.03) |
| 12/01/2022 - | Median | 0 | 0 | 0 | 0.01 | 0.01 | 0.01 | 0.01 |
| 2/28/23 | (Q1; Q3) | (0; 0) | (0; 0.01) | (0; 0.01) | (0; 0.02) | (0; 0.02) | (0; 0.03) | (0; 0.03) |

**Note: MAE: day x prediction: County level MAE for x days ahead prediction; Q1: First Quartile; Q3: Third Quartile.**

**Table A16. County-Level Mean Absolute Error (MAE) and interquartile range (IQR) for weekly prediction (Persistence model).**

| | | MAE: week 1 prediction | MAE: week 2 prediction | MAE: week 3 prediction | MAE: week 4 prediction | MAE: week 5 prediction |
|---|---|---|---|---|---|---|
| **cases** | | | | | | |
| 12/01/2021 - | Median | 138.36 | 267.22 | 374.63 | 454.5 | 515.66 |
| 3/31/22 | (Q1; Q3) | (67.94; 359.6) | (126.11; 702.9) | (176.18; 984.11) | (213.96; 1186.97) | (239.4; 1347.23) |
| 05/01/2022 - | Median | 30.48 | 38.22 | 48.99 | 56.26 | 61.92 |
| 10/31/22 | (Q1; Q3) | (11.25; 49.34) | (16.04; 74.79) | (20.59; 92.61) | (22.24; 120.7) | (24.54; 141.82) |
| 12/01/2022 - | Median | 19.38 | 26.91 | 36.11 | 41.27 | 46.08 |
| 2/28/23 | (Q1; Q3) | (10.13; 46.48) | (15.42; 60.2) | (16.82; 81.82) | (19.99; 95.48) | (22.2; 109.48) |
| **deaths** | | | | | | |
| 12/01/2021 - | Median | 0.5 | 0.8 | 1.11 | 1.32 | 1.5 |
| 3/31/22 | (Q1; Q3) | (0.19; 0.94) | (0.36; 1.64) | (0.5; 2.33) | (0.64; 2.93) | (0.72; 3.58) |
| 05/01/2022 - | Median | 0.1 | 0.2 | 0.29 | 0.4 | 0.49 |
| 10/31/22 | (Q1; Q3) | (0.05; 0.22) | (0.1; 0.4) | (0.16; 0.58) | (0.21; 0.76) | (0.28; 0.95) |
| 12/01/2022 - | Median | 0.06 | 0.12 | 0.16 | 0.22 | 0.26 |
| 2/28/23 | (Q1; Q3) | (0.03; 0.2) | (0.05; 0.38) | (0.08; 0.5) | (0.11; 0.57) | (0.13; 0.63) |

**Note: MAE: week x prediction: County level MAE for x weeks ahead prediction; Q1: First Quartile; Q3: Third Quartile.**
